# Social Isolation and All-Cause and Cause-Specific Mortality in the UK Biobank: The Modifying Role of Non-Commute Transportation Mode

**DOI:** 10.64898/2026.09.21.26363514

**Authors:** Emily Diem Hum, Yacine Lapointe, Jill Baumgartner, Kevin Manaugh, Hiroshi Mamiya

**Affiliations:** Department of Epidemiology, Biostatistics, and Occupational Health, McGill University., Suite 1200, 2100 McGill College, Montreal, QC, Canada, H3A 1G1; Department of Equity, Ethics, and Policy, McGill University. Suite 1200, 2100 McGill College, Montreal, QC, Canada, H3A 1G1; Department of Geography, McGill University. Room 305, 805 Sherbrooke Street West, Montreal, Quebec H3A 0B9; Bieler School of Environment, McGill University. 3534 University Montreal, Quebec H3A 2A7

**Author notes:** Corresponding author, Hiroshi Mamiya, Suite 1200, 2100 McGill College, Montreal, Quebec, Canada H3A 1G1.

**Keywords:** Social isolation, transportation, mortality, aging

## Abstract

**Background:** Social isolation (SI) is an established risk factor for chronic disease and mortality. How people travel, including the modes of transportation they use, may shape opportunities for social interaction and influence the impacts of SI. This study assessed heterogeneity of the association between SI and mortality across primary modes of non-commute transportation.

**Methods:** We analyzed cohort data from the UK Biobank, following participants aged 40-69 years from 2006 to 2022. SI at baseline was derived from three items and dichotomized.

Cox models were used to estimate the association between SI and mortality, adjusting for socioeconomic factors, health behaviours, and comorbidities. Effect modification by participants’ primary mode of non-commute transportation was assessed using interaction terms with SI.

**Results:** There were 490,114 participants with complete baseline exposure data. SI was associated with increased hazard of all-cause (Hazard Ratio [HR] = 1.34, 95% Confidence Interval [CI]: 1.31-1.38) and cardiovascular mortality (HR = 1.49, CI: 1.41-1.57). The association with cardiovascular mortality was stronger among participants who only used public transport compared with those who only used a car (interaction HR = 1.17, CI: 1.00-1.37). In contrast, using a car alongside active or public transport, compared to using a car alone, attenuated the association between SI and all-cause mortality (interaction HR = 0.93, CI: 0.87-0.99).

**Conclusion:** Mixed transport, including a car, showed the greatest reduction in the association between SI and all-cause mortality. Our findings motivate further exploration of the role of transportation modes in enhancing social connection, mobility, and health.

## Introduction

Social isolation (SI) is an emerging public health priority worldwide.^1^ SI refers to the objective absence or limitation of social contacts and relationships and is conceptually distinct from loneliness, which captures the subjective experience of feeling alone.^2^

Although related, these constructs do not fully overlap. Individuals may have limited social contact without feeling lonely, and, conversely, can feel lonely even with social contact.

A growing body of evidence links SI with all-cause and cause-specific mortality from cancer and cardiovascular disease (CVD).^3,4^ Several mechanisms have been proposed. First, SI may influence physiological processes such as endocrine signaling, leading to adverse changes in heart rate, blood pressure, and vessel repair.^5^ Additionally, SI can lead to changes in brain structure and processes, sympathetic neural tone, cortisol secretion, and impaired immune function.^5^ Second, SI is associated with adverse health behaviors including smoking, alcohol consumption, overeating, and poor sleep, which may act as mediators between SI and adverse health outcomes.^5^ Third, SI is associated with an increased risk of depression and cognitive decline in later life, which, in turn, may increase risk of mortality.^4^ Finally, socially isolated individuals may be less likely to access preventive and acute health care, increasing their risk of poor health outcomes.^4^

Most interventions targeting SI have focused on individual or small-group approaches, such as social prescribing, group activities, and peer support.^6–8^ While SI is indeed experienced at the individual level, it is also influenced by the social, structural, and environmental conditions that shape opportunities for social connection. There is increasing interest in identifying environmental interventions to mitigate SI.^9,10^

Daily transportation modes, which are strongly shaped by modifiable features of the built environment, may influence SI by enhancing mobility to meet people, and themselves acting as a third place for social interaction.^11,12^ In particular, non-commute travel may facilitate engagement in social and recreational activities, especially in older adults.^11^ Evaluating whether transportation modes modify the health effects of SI could help inform transportation-based interventions to promote social connection, health, and mobility. Yet, no studies have addressed this question to date. In this study, we examine whether non-commute transportation mode modifies the association between SI and all-cause and cause-specific mortality using UK Biobank data. Given the conceptual overlap but distinct measurement, we also assessed loneliness in sensitivity analysis.

## Methods

This study used longitudinal data from the UK Biobank, a prospective cohort of over 500,000 adults aged 40-69 years, followed from 2006 to 2022 across 22 centers in the U.K.^13^ Participants who withdrew consent (∼1,200) or were missing baseline SI data (n=10,695) were excluded, yielding an analytic sample of 490,114 participants (**Appendix Figure 1**).^14^

SI was assessed via self-administered touchscreen questionnaires between 2006 and 2010. Using previous methods, we constructed a composite index based on three indicators: 1) infrequent contact with family and friends (<= once per month), 2) no weekly participation in social activities, and 3) living alone.^15^ Participants meeting at least two criteria were classified as socially isolated.^15^ Loneliness was assessed in sensitivity analysis using a single-item dichotomous measure (“Do you often feel lonely?”).

UK Biobank recorded time and cause of death through linkages with national death registries. Events of interest included all-cause mortality and cause-specific mortality from three conditions previously associated with SI (cancer, CVD, and chronic respiratory conditions).^16,17^ Cause of death was ascertained using ICD-10 codes (**Appendix Table 1**).

We examined usual mode of transportation for non-commute travel as a potential effect modifier. Participants reported on the most common transport modes used in the past four weeks (excluding commuting), with the option to select multiple modes. Because frequency and duration were not captured, we categorized responses as: active transport only (walking or cycling), public transport only, mixed-without-car, mixed-with-car, and car-only (**Appendix Text 1**). We focused on non-commute travel given its relevance to discretionary activities and social participation, particularly for an older cohort where a proportion (43%) were retired or unemployed at baseline.

Covariate selection was informed by directed acyclic graphs (DAGs) based on previous literature investigating the association between SI and mortality (**Appendix Figures 2-3**).^4,15,17,18^ See **Appendix Text 1** for model specifications and covariate categories. The primary model adjusted for sociodemographic and contextual factors that may plausibly act as confounders, including age, biological sex, ethnicity, Townsend deprivation index, annual household income, a combined indicator of employment status and commuting transportation mode, population density, and survey assessment month.

Chronic health conditions^5,19^ and health behaviors such as smoking,^20^ alcohol consumption,^21^ physical activity,^22^ sleep,^23,24^ and diet^25^ were not included in the primary model, as they may lie on the causal pathway between SI and mortality. However, chronic conditions can limit social participation and impact the quality of relationships.^26,27^

Smoking may isolate individuals due to stigma and thereby increase SI.^20,28^ Alcohol consumption, dietary patterns, and physical activity in group settings may facilitate social interaction,^25,29,30^ while poor sleep may negatively affect social relationships.^31^ Thus, we examined their inclusion in sequential models to assess sensitivity to potential overadjustment.

We first described participants’ baseline characteristics by mortality status using counts and proportions for categorical variables and medians (interquartile ranges) for continuous variables. Follow-up time was calculated as the time in months from baseline assessment to date of death or administrative censoring (December 19, 2022), whichever occurred first.

We constructed Kaplan-Meier survival curves and fit Cox proportional hazards models for SI as a predictor of mortality outcomes. Separate models were fit for all-cause mortality and each cause-specific outcome. The primary model adjusted for sociodemographic-, time-, and place-related confounders identified *a priori*. Hazard ratios (HRs) and 95% confidence intervals (CIs) were reported. To assess effect measure modification, we included interaction terms between SI and categories of non-commute transportation mode, using car-only as the reference group. We reported stratum-specific estimates and interaction terms, with statistical evidence for interactions evaluated using Wald tests.^32^ To aid interpretation, we examined both relative differences over the follow-up period (multiplicative interaction HRs) and absolute differences in predicted 10-year absolute risk (additive scale, detailed methods in **Appendix Text 2**).

The proportional hazards assumption was assessed using visual inspection of scaled Schoenfeld residual plots. No deviations were observed. Participants with missing baseline covariates were excluded from the complete-case analysis. We compared distributions of key variables between included and excluded participants to assess potential selection bias.

We conducted a series of pre-specified sensitivity analyses to assess the robustness of our findings. First, we sequentially adjusted for variables that may lie on the causal pathway, including smoking (Model 2), additional health behaviors (Model 3), and chronic conditions (Model 4), to evaluate the extent to which estimates were sensitive to potential overadjustment. Commuting transportation mode was the most common missing variable, with 4.5% missingness (n=21,871/490,114). To assess potential selection bias from excluding these participants, we fit a model adjusting for main covariates without commute transportation mode. Second, we repeated all analyses treating loneliness as the exposure. Third, for cause-specific mortality outcomes, we conducted competing risk analyses using Fine-Gray subdistribution hazard models, treating deaths from other causes as competing events.^33^ These models estimate subdistribution HRs, which reflect the cumulative incidence function. Fourth, we excluded deaths occurring within two years from baseline to reduce potential reverse causation. Fifth, we restricted analyses to participants reporting zero hours of driving per week to examine associations among non-drivers. Sixth, we restricted analyses to participants residing in urban areas, given potential differences in transportation infrastructure. Seventh, we excluded participants who selected multiple transportation modes. Finally, we conducted sex-stratified analysis to examine heterogeneity.

Analyses were conducted in R (4.5.2),^34^ using the *survival* (3.7-0),^35^ *fastcmprsk* (1.24.10),^36^ and *riskRegression* (2025.09.17)^37^ packages. Software code is available upon request. The study was approved by the Faculty of Medicine Institutional Review Board (A08-M55-24A). We provided the STROBE checklist (**Appendix Text 3**).

## Results

Over the study period (median follow-up approximately 165 months), 42,913 deaths occurred **(Table 1)**. Participants who died were more likely to be socially isolated at baseline than those who remained alive (19% versus 14%, respectively). They were also more likely to be male (59% versus 44%), have no formal educational qualifications (31% versus 16%), and have an annual household income less than £18,000 (34% versus 18%). Most participants reported primarily using transportation modes involving a car (78% across car-only and mixed-transport-with-car categories) (**Appendix Figure 4**).

**Table 1.**
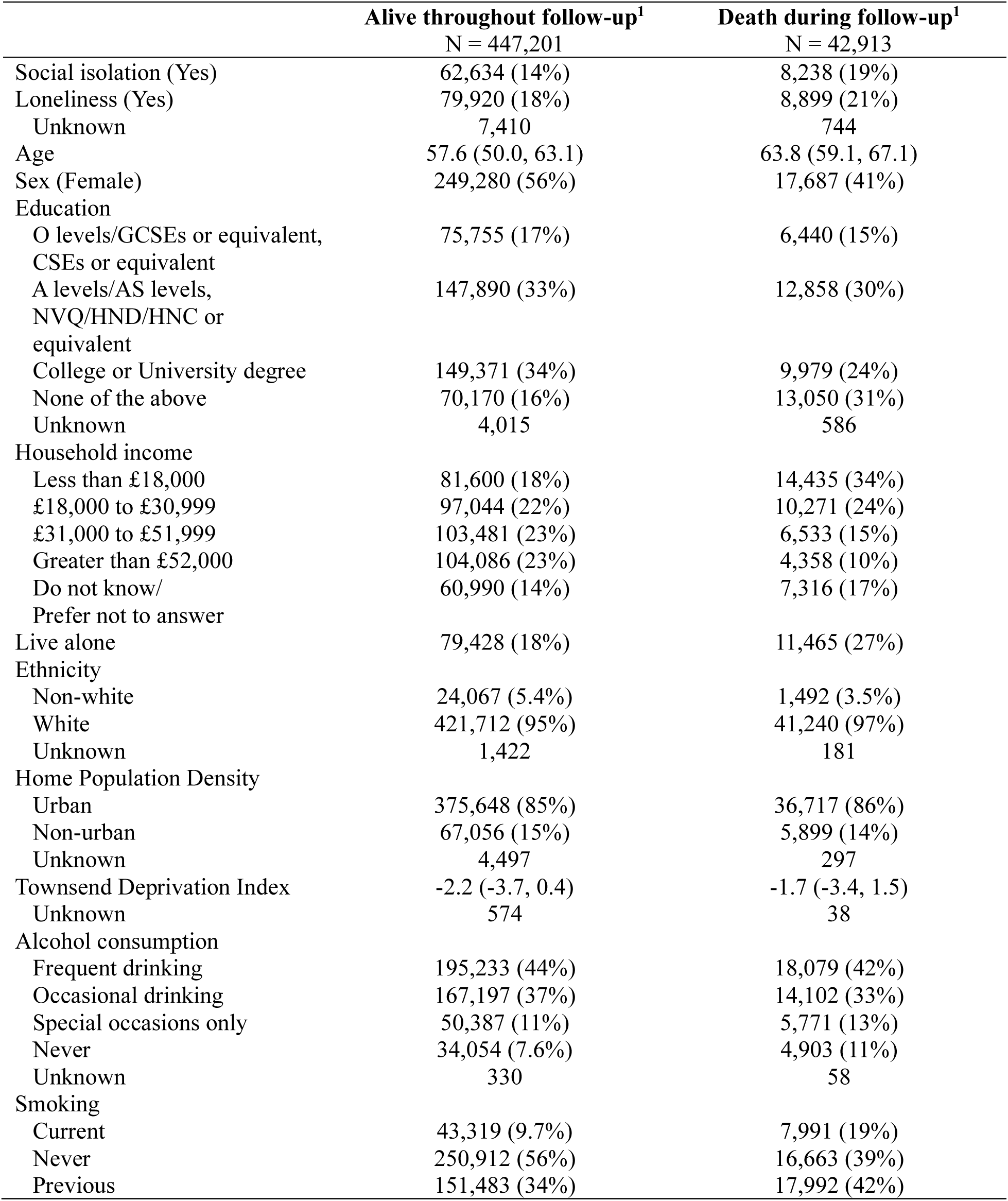
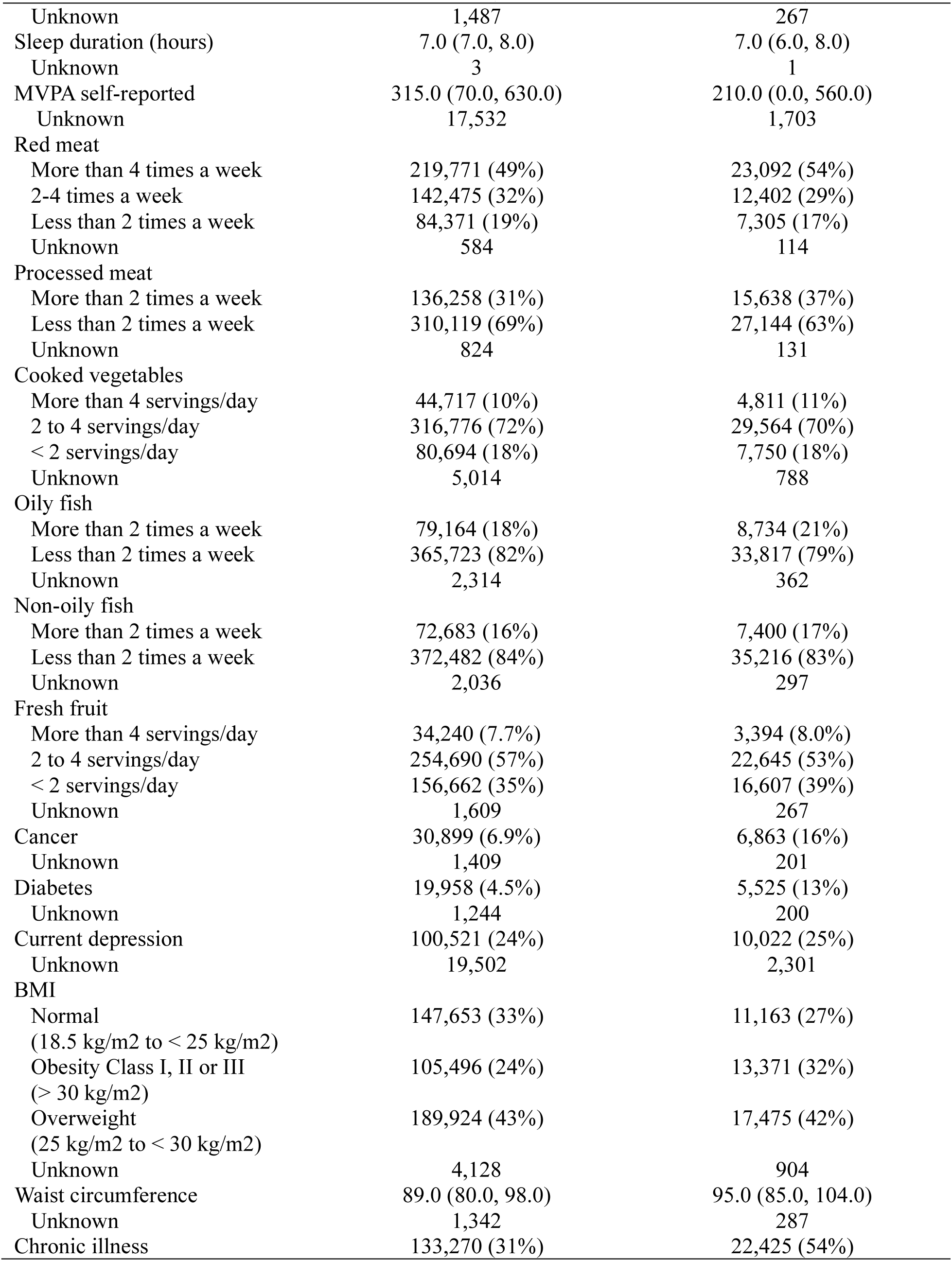
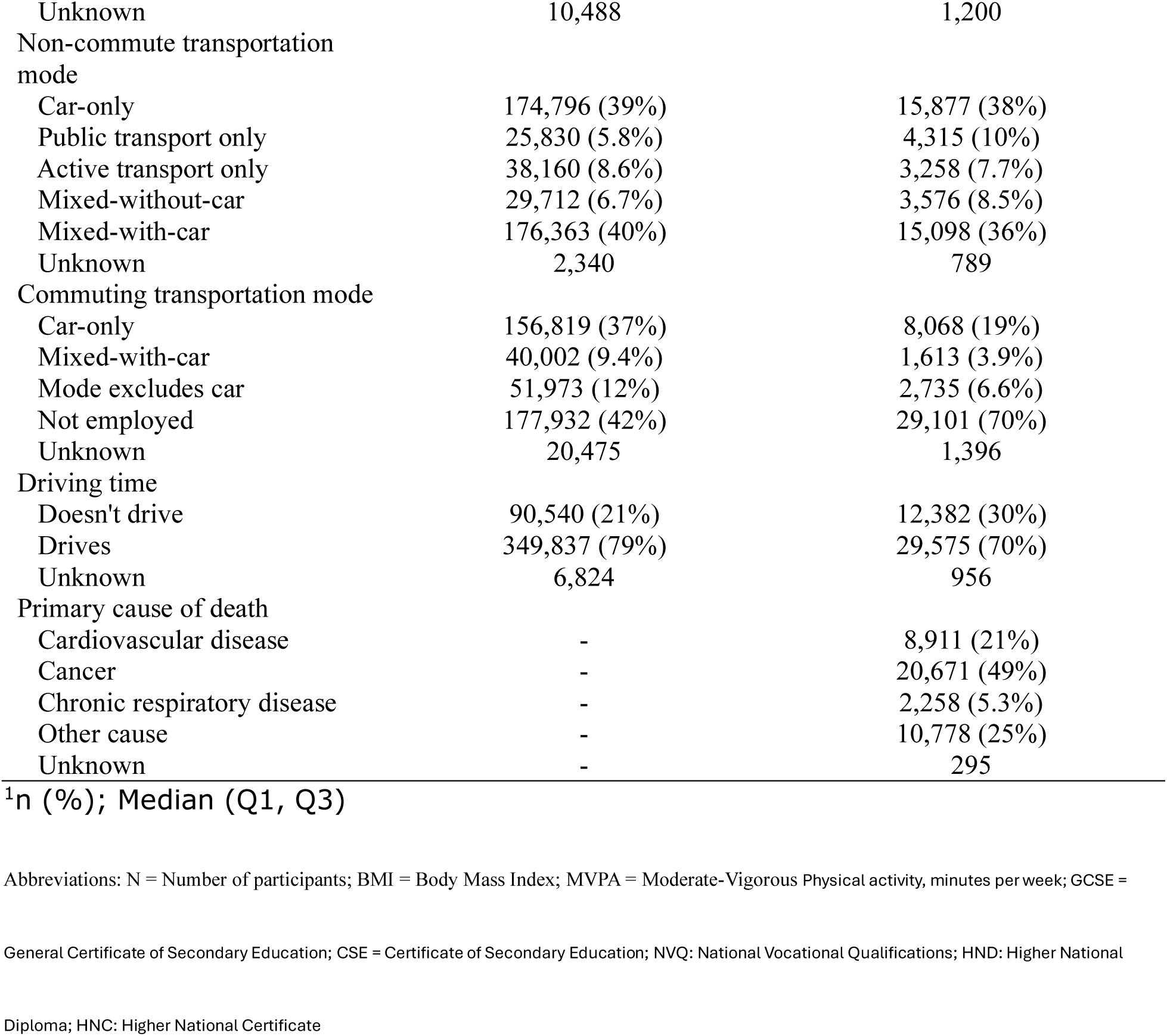
Distribution of baseline participant characteristics by death status.

In unadjusted analyses, SI was associated with higher mortality across all outcomes (**Figure 1**), consistent with Kaplan-Meier curves showing lower survival probabilities among socially isolated participants (**Appendix Figure 5**). After adjustment for sociodemographic covariates, month of assessment, and population density, SI remained associated with increased hazards of all-cause mortality (HR = 1.34, CI: 1.31-1.38), and cause-specific mortality from CVD (HR = 1.49, CI: 1.41-1.57), cancer (HR = 1.22, CI: 1.18-1.27), and chronic respiratory conditions (HR = 1.65, CI: 1.49-1.83). In interaction models, the association between SI and all-cause mortality was weaker among participants using mixed-modes (including a car) compared with car-only (interaction HR = 0.93, CI: 0.87-0.99) (Figure 2). Estimates for other modes were close to the null and did not suggest clear patterns of effect modification. Point estimates ranged from modestly protective, for active transport only, to slightly increasing the SI-mortality association, for public transport only and mixed-transport-without-car, though their CIs included the null.

**Figure 1.** Hazard ratios and 95% confidence intervals for the association between social isolation and mortality outcomes, without interactions between non-commute transportation mode and social isolation

**Figure 2.**
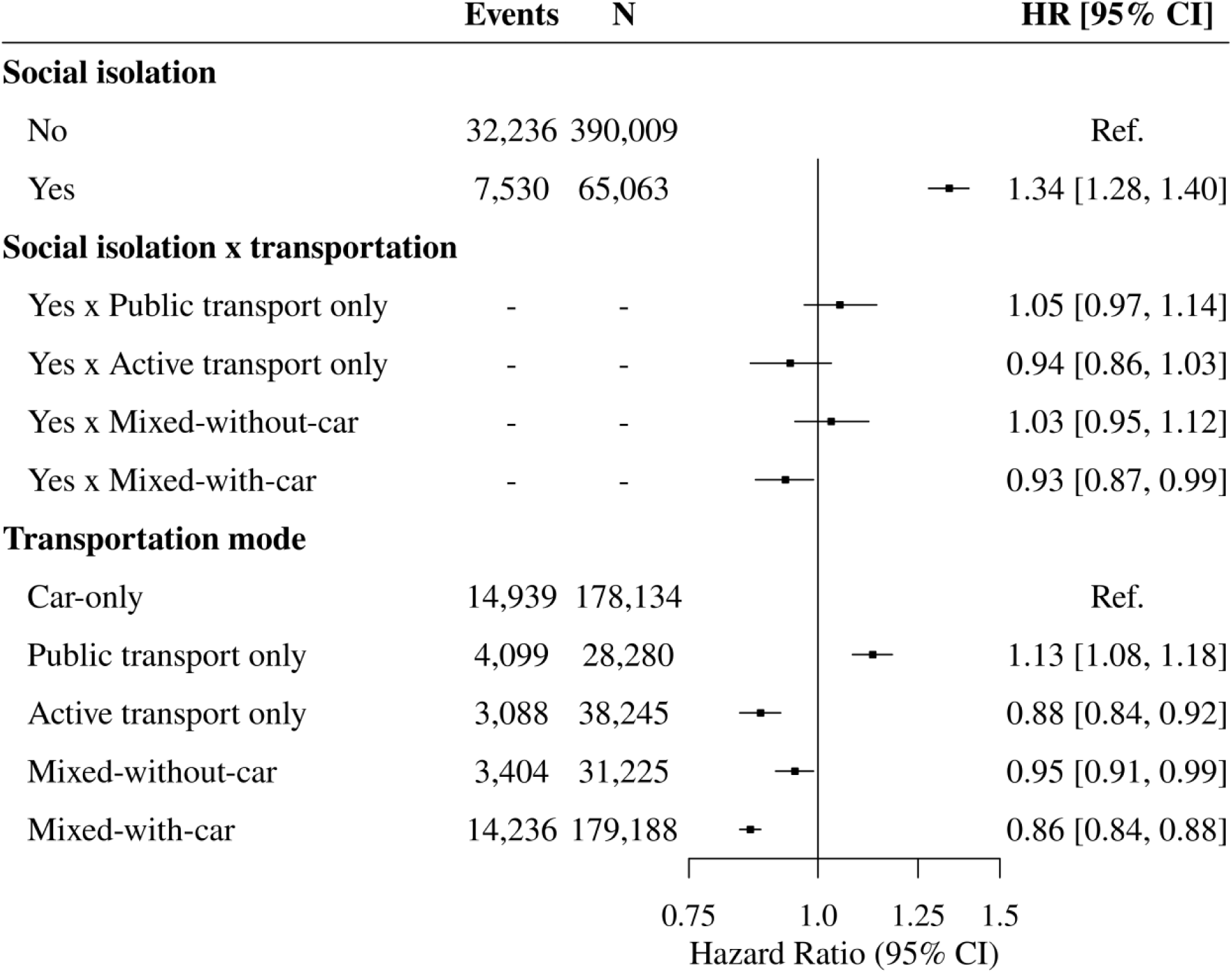
Association of social isolation with all-cause mortality by non-commute transport subgroup, using car-only as the reference category Model adjusted for age, sex, ethnicity, education, Townsend deprivation index, annual household income, month of assessment, commute transportation mode, and population density. Abbreviations: No. deaths = Number of deaths; N = Total number of participants; HR = Hazard ratio; CI = Confidence interval; Ref. = Reference category. Arrowheads indicate confidence intervals extending beyond the displayed x-axis range.

For CVD mortality, there was some evidence that the SI-mortality association was larger among participants who used public transport only relative to car-only users (interaction HR = 1.17, CI: 1.00-1.37) (Figure 3). Patterns for other transportation modes were consistent with those for all-cause mortality, though CIs included the null value, suggesting limited effect modification. We found no evidence of effect measure modification by transport mode for cancer or chronic respiratory mortality (**Appendix Tables 4-5**).

**Figure 3.**
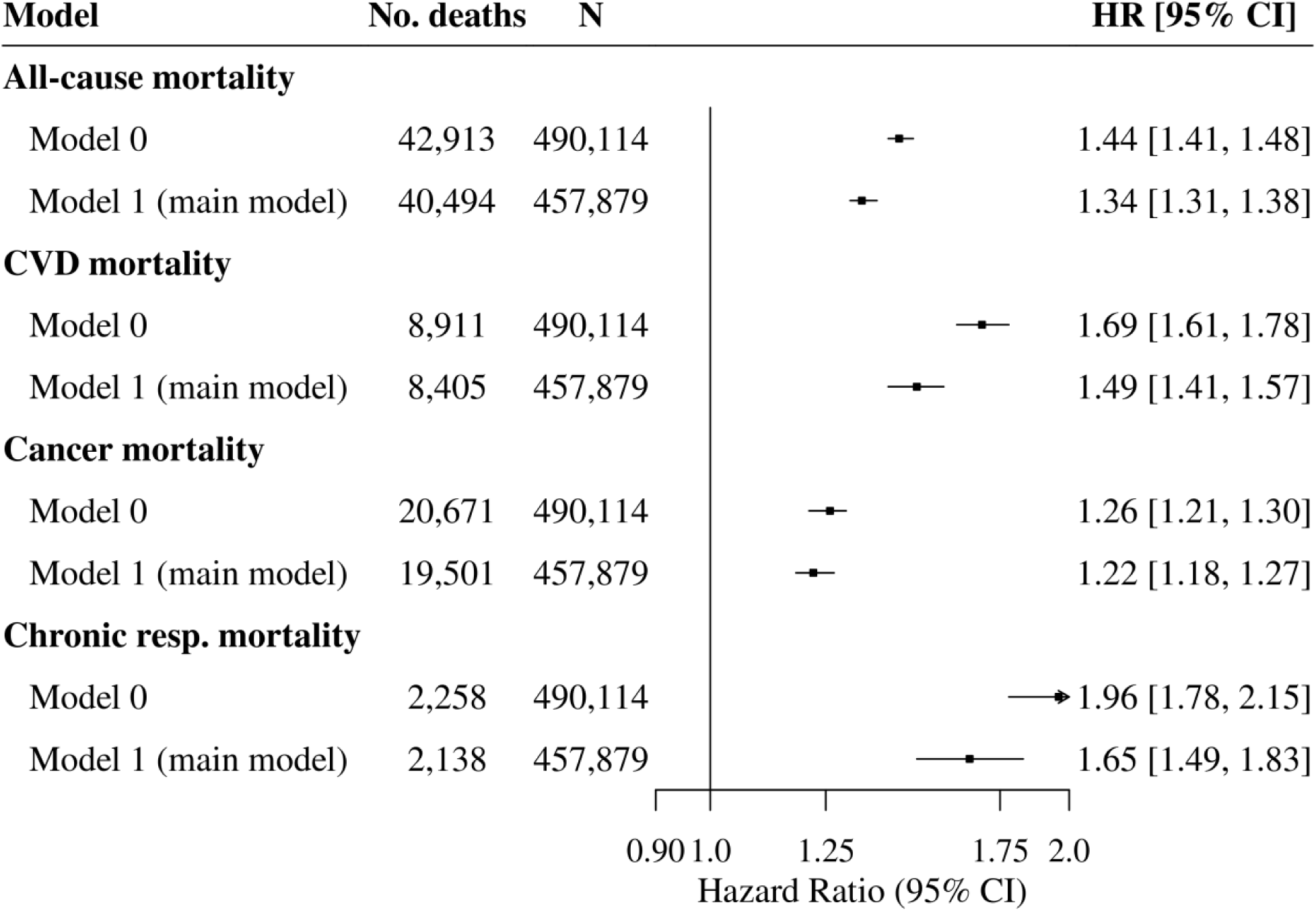
Hazard ratios and 95% confidence intervals for the association between social isolation and mortality outcomes, without interactions between non-commute transportation mode and social isolation Model 0 unadjusted. Model 1 adjusted for age, sex, ethnicity, education, Townsend deprivation index, annual household income, commute transportation mode, month of assessment, and population density. Abbreviations: Chronic resp. mortality = Chronic respiratory mortality; No. deaths = Number of deaths; N = Total number of participants; HR = Hazard ratio; CI = Confidence interval. Arrowheads indicate confidence intervals extending beyond the displayed x-axis range.

**Figure 3.**
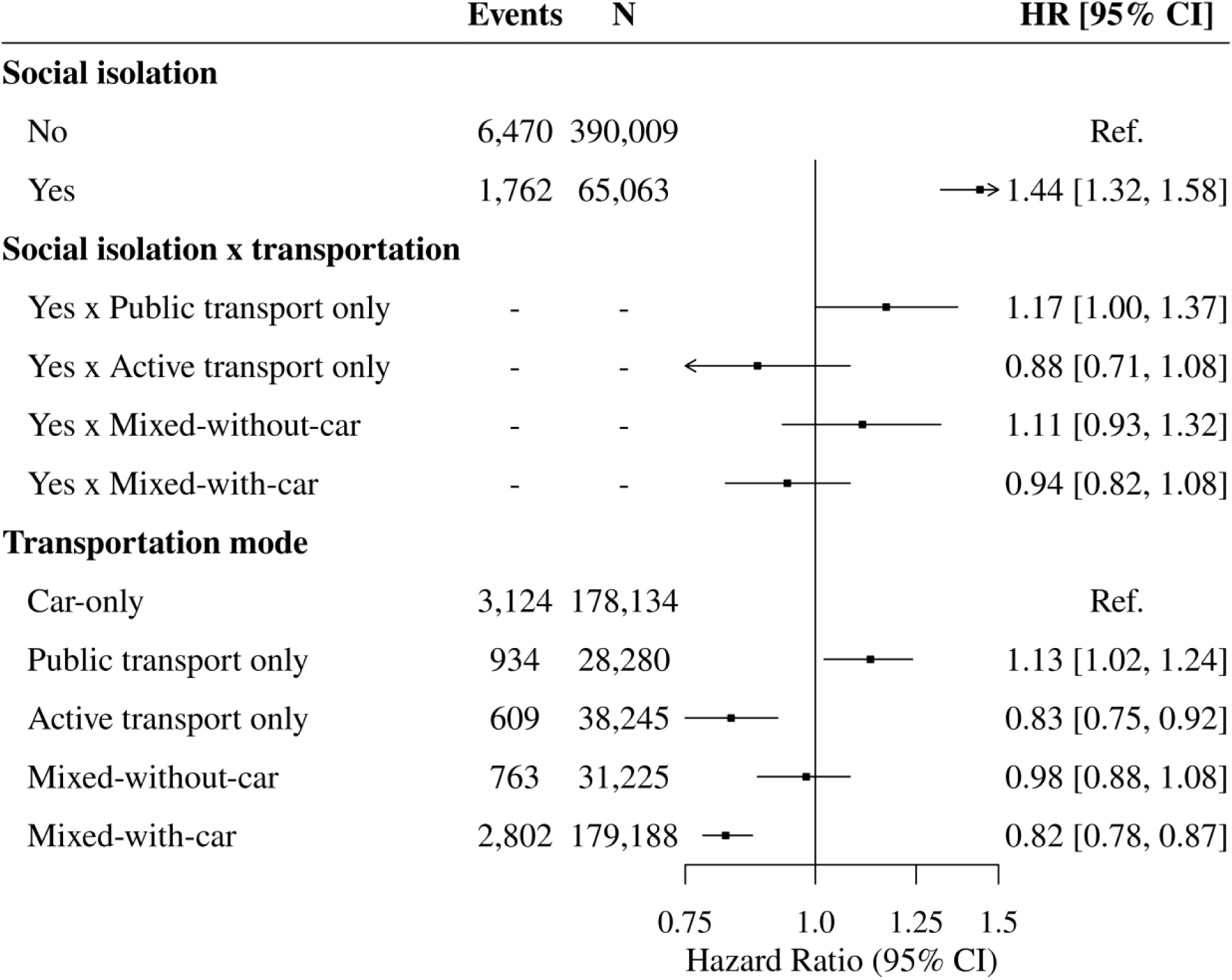
Association of social isolation with cardiovascular mortality by non-commute transport subgroup, using car-only as the reference category Model adjusted for age, sex, ethnicity, education, Townsend deprivation index, annual household income, month of assessment, commute transportation mode, and population density. Abbreviations: No. deaths = Number of deaths; N = Total number of participants; HR = Hazard ratio; CI = Confidence interval; Ref. = Reference category. Arrowheads indicate confidence intervals extending beyond the displayed x-axis range.

Additive effect modification results were consistent with findings from the multiplicative scale (**Appendix Figures 6-7 and Table 6**). For all-cause mortality, the joint effect of SI and using mixed-modes (including a car), compared to car-use alone, corresponded to approximately 305 fewer deaths per 100,000 persons attributable to the interaction between the two exposures, beyond the sum of their individual effects. In contrast, approximately 288 and 106 excess all-cause and CVD deaths per 100,000 persons, respectively, were associated with the joint effect of SI and public transportation use alone. Although several additive interaction estimates were statistically significant, these findings should be interpreted cautiously, as estimates were derived from model-based predictions and corresponding standard errors rather than resampling-based methods (**Appendix Text 2**).

Results from sensitivity analyses are presented in **Appendix Figures 8-9 and Tables 7-26**.

Additional adjustment for health behaviors and chronic conditions attenuated the observed associations between SI and mortality, with HRs decreasing by up to 0.43, but did not eliminate them. Interaction estimates for mixed-mode transportation (including a car) and SI in relation to all-cause mortality changed minimally across models and suggested effect modification patterns in line with our main findings. Precision was comparable across models, although CIs shifted to include the null in Models 3 and 4. For CVD mortality, the interaction between public transportation and SI decreased modestly in magnitude across Models 2-4, with little change in precision; however, CIs included the null following these adjustments.

Analyses using loneliness as the exposure yielded similar overall patterns, with increased hazards of all-cause, CVD, cancer, and chronic respiratory mortality. The interaction between loneliness and active transportation on CVD mortality was newly significant, indicating that the association between loneliness and CVD mortality was 23% stronger for active transport users compared to car-only users. This remained significant when adjusting for health behaviors and comorbidities (Model 4). A similar pattern was observed between loneliness and mixed transportation without a car after adjusting for Model 4 covariates.

Results from competing risk models, removing commute transportation from main covariates, exclusion of early deaths, restriction to urban populations, and restricting to participants who only selected one mode were broadly consistent with the main findings, although interaction effects were generally attenuated and often no longer statistically distinguishable from the null.

Among non-drivers, the modifying effect of mixed-modes-with-car for all-cause mortality was strongly attenuated towards the null, where car use presumably involved relying on lifts from others or ridesharing. The direction of interaction for other transportation modes in both all-cause and CVD mortality was consistent with our main findings, though CIs widened to include the null. In loneliness models, public transport use vs. car-only use became protective against all-cause mortality attributed to loneliness. Active transportation had a similar protective effect against loneliness-attributed chronic respiratory mortality.

In sex-stratified analysis, the protective interaction between mixed-transport-with-car and SI on all-cause mortality, relative to car-only, was statistically significant among males, but not among females. Similarly, there was some evidence of the harmful interaction between public transport only and SI for CVD mortality among males, but not females.

## Discussion

To our knowledge, this is the first study to assess the role of non-commute transportation in modifying the association between SI and mortality. SI was associated with higher all-cause, CVD, cancer, and chronic respiratory mortality. The association with CVD mortality was stronger among participants who primarily used public transport compared with car-only users. In contrast, using a car alongside active or public transport attenuated the association between SI and all-cause mortality. These findings suggest a gradient of vulnerability to SI-attributed mortality, wherein mixed-mode with a car was most protective, followed by car-only, and finally, modes without a car.

Previous literature suggests that public transportation plays an important role in supporting mobility and social connection among aging adults.^38^ However, we found that car use mitigated SI-attributed mortality compared to public transportation alone. A plausible explanation is that car use affords the greatest mobility freedom among the examined transportation modes.^39^ Relying on public transportation infrastructure may be insufficient to compensate for the loss of mobility when using a car is not possible. Among socially isolated individuals, these limitations, compounded with reduced social support, can limit access to health care services,^40^ increase stress, and lead to adverse mental and physical health.^41^ However, the possible benefits of active and public transportation are supported by our second finding: using mixed-modes with a car, compared to a car alone, reduced SI-attributed all-cause mortality. Public and active transportation are both associated with positive social, mental, and physical health benefits.^42–46^ Additionally, mixed-modes, as opposed to relying on single-modes, may increase freedom and travel flexibility.^47^ Thus, increasing the uptake of active and public transportation among car users may be a promising point of intervention to reduce the adverse impact of SI.

Adjusting for potential mediators attenuated estimates, potentially due to overadjustment bias (adjusting for mediators rather than confounders).^48,49^ Results must therefore be interpreted with caution. Overall, sensitivity analyses were consistent with our main findings. Loneliness analyses suggested that car use was most protective against loneliness-attributed mortality, while among non-drivers, using public or active transportation (vs. relying on rides from others) was more protective. This finding aligns with a recent systematic review, which found that public transportation is especially important for reducing loneliness when driving is not an option.^11^ The stronger protective effect of car use among males aligns with existing literature suggesting that men often place greater value on driving,^11^ which may increase their risk of adverse outcomes when they cannot drive. Due to data limitations, we were unable to analyze gender, rather than biological sex. Nevertheless, our study provides compelling evidence for further exploration of gender differences in transportation-related interventions.

The strength of this study lies in its novel investigation of effect modification by transportation mode, enabled by the large sample size of the UK Biobank. The study presents some limitations. First, the UK Biobank comprises a highly educated, predominantly White cohort, which may limit generalizability. However, previous work found minimal differences between unweighted and post-stratified UK Biobank estimates, suggesting reduced risk of selection bias.^50^ Second, although adapted from the Berkman-Syme Social Network Index,^51^ UK Biobank’s SI index did not capture components used elsewhere (e.g., marital status). However, previous studies have reported comparable estimates between the study’s index and widely used scales.^52^ Third, loneliness was measured using a single-item question, which, compared to validated multi-item instruments,^17^ is more prone to social desirability bias and potential differential misclassification with an unpredictable direction of bias.^53^ Loneliness findings were therefore examined in sensitivity analysis and should be interpreted with caution. Fourth, SI and covariates were only assessed at baseline, as ∼80% of participants did not complete follow-up questionnaires. Among participants who were followed, a relatively small proportion transitioned into (8.7%) and out of (8.0%) SI. Key covariates have also been shown to be stable over time in the cohort.^54^ However, our inability to investigate time-varying effects remains a limitation. Fifth, reporting of non-commute transportation modes may have been inconsistent, as participants could select multiple modes without guidance on frequency of use. This may have introduced non-differential misclassification, which is expected to attenuate interaction estimates towards the null. Sensitivity analyses restricted to single-mode users yielded results consistent with our main findings. Sixth, we conducted complete-case analysis, excluding participants with missing SI (2.1%, n=10,709/500,823) and covariates (7.0%, n=35,042/500,823), which introduced potential selection bias. We did not conduct multiple imputation as there was less than 10% missingness.^54^ Individuals missing SI were moderately more socioeconomically disadvantaged and less likely to use a car compared to those included for analysis (**Appendix Tables 27-28**), which may have attenuated observed associations. Conversely, participants missing covariates did not differ meaningfully from included participants.

Sensitivity analyses omitting commute mode, the variable with the largest missingness, were consistent with our main findings. Finally, as in all observational studies, residual confounding poses a risk to the validity of our findings.

Future work could explore the modifying role of transportation modes in different contexts, as well as other dimensions of transportation (e.g. commuting, transport accessibility).

Furthermore, research incorporating time-varying exposures and validated measures of loneliness may be warranted. Overall, mixed-modes with a car appeared to be more protective than a car alone, which, in turn, was more protective than not using a car at all for mitigating SI-attributed mortality. Since accessing a car is not always possible, especially among older adults,^56^ our study motivates further efforts to improve public and active transportation infrastructure in ways that support social connection, mobility, and health.

## Data Availability

This study used data from the UK Biobank (https://www.ukbiobank.ac.uk). The data are not openly available. They can be accessed by approved researchers through an application to UK Biobank, which provides credentialed access to the data (UK Biobank Application Number 45551).

## Acknowledgements

This research was conducted using UK Biobank data under application No 45551.

## Data availability statement

UK Biobank data are available for research upon approval from the UK Biobank Access Management System.

## Supplementary materials for “Social Isolation and Mortality in the UK Biobank: The Modifying Role of Non-Commute Transportation mode Mode”

## Supplementary materials

**Appendix Figure 1.**
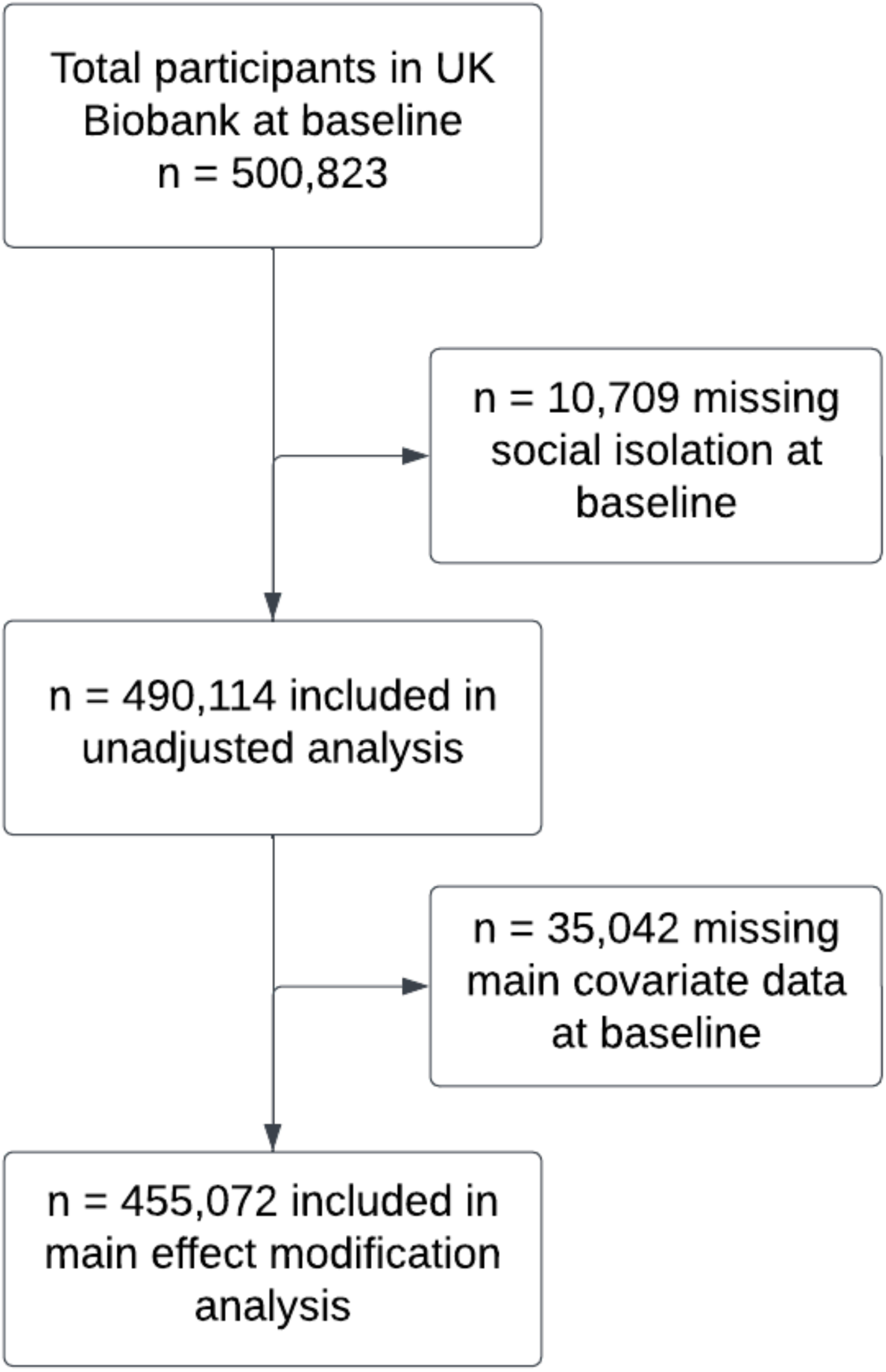
Participant flowchart

**Appendix Table 1.**
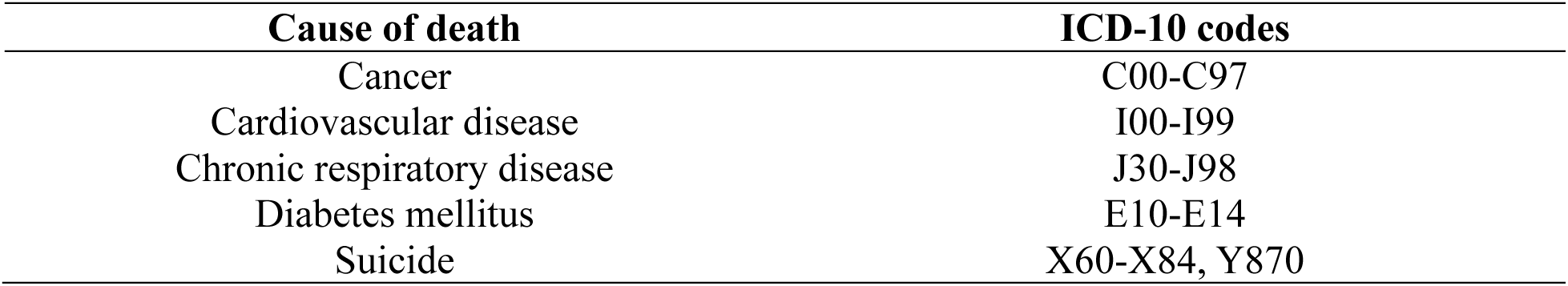
Cause of death coding.

| Cause of death | ICD-10 codes |
| --- | --- |
| Cancer | C00-C97 |
| Cardiovascular disease | I00-I99 |
| Chronic respiratory disease | J30-J98 |
| Diabetes mellitus | E10-E14 |
| Suicide | X60-X84, Y870 |

**Appendix Figure 2.**
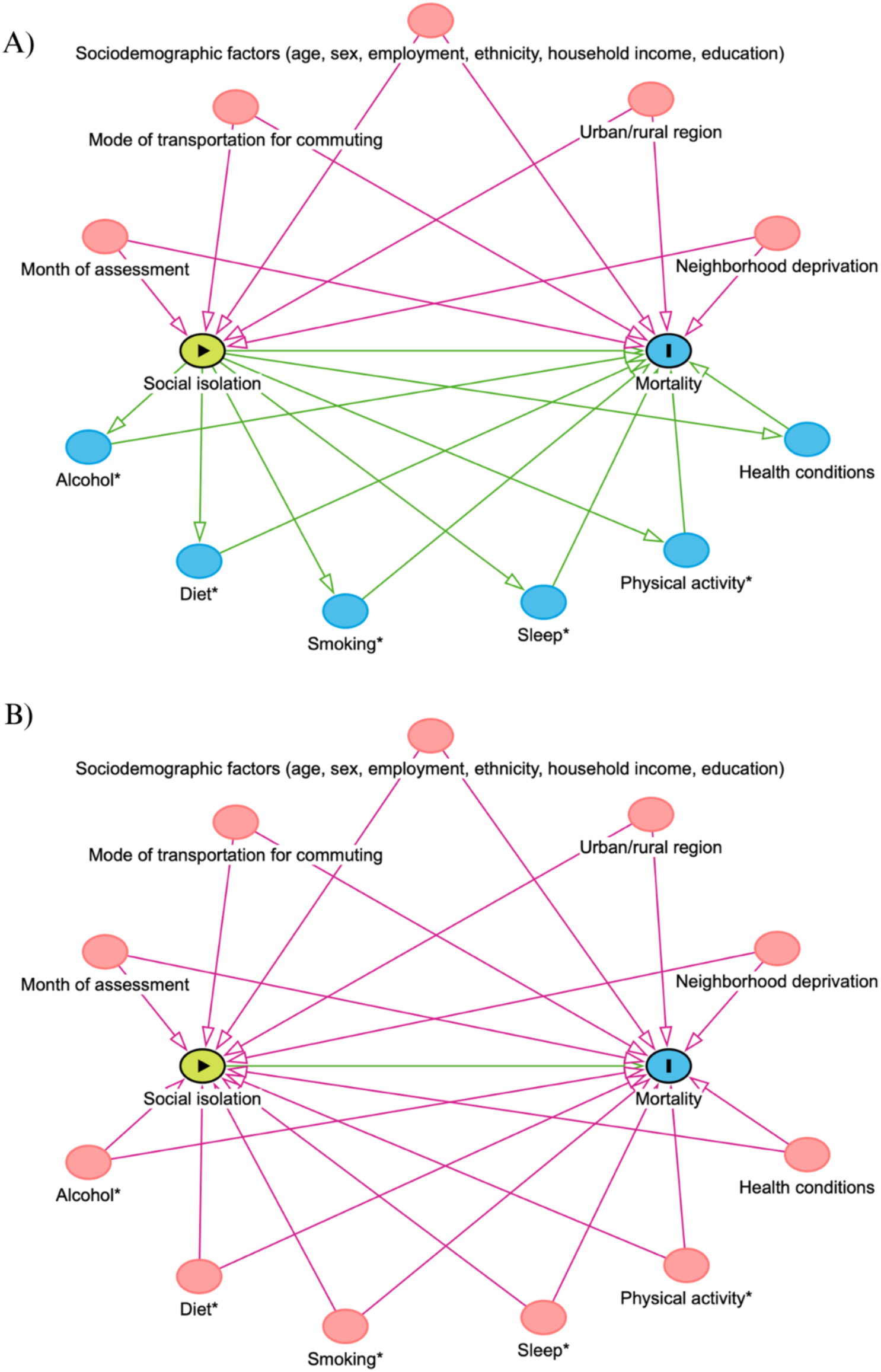
Directed acyclic graphs illustrating confounders in the relationship between social isolation and mortality

Figure created using DAGitty. Potential confounders are illustrated in pink. Potential mediating pathways are illustrated in green.

Social isolation can lead to changes in health behaviors and increased risk of chronic disease.^1–5^ These factors may therefore be mediators on the causal pathway between social isolation and mortality (Figure 2a. A). However, health behaviors, including smoking, alcohol, diet, sleep, and physical activity, as well as health conditions, have both been causally associated with social isolation and mortality (Figure 2a. B). Smoking may isolate individuals due to stigma and thereby increase risk of social isolation.^1,6^ Alcohol consumption, dietary patterns, and physical activity in group settings may facilitate social interaction,^4,7,8^ while poor sleep may negatively affect social relationships.^9^ Finaly, chronic conditions may limit participation in social activities and impact the quality of social relationships.^10,11^

---

**Appendix Figure 3.**
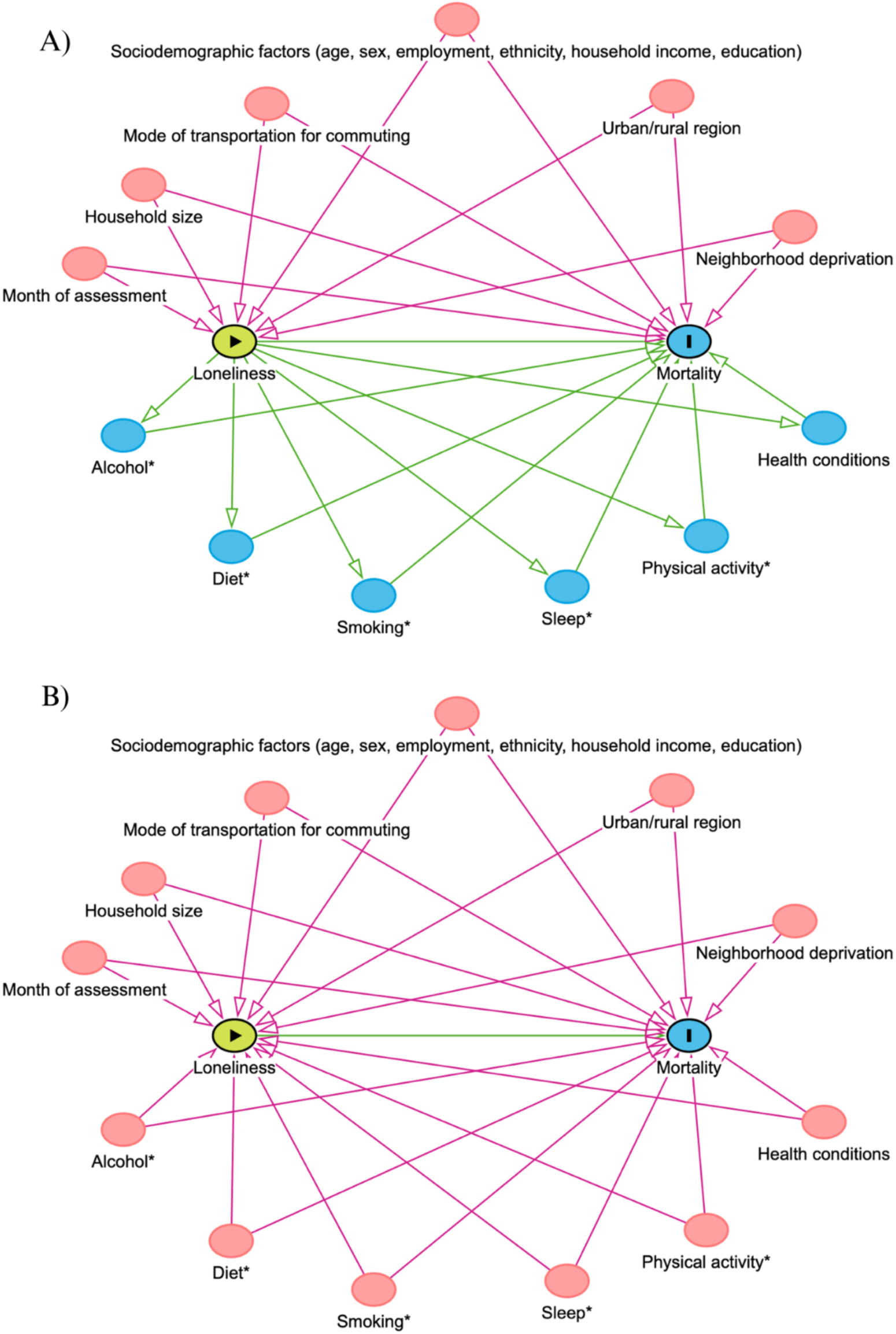
Directed acyclic graphs illustrating confounders in the relationship between loneliness and mortality

Figure created using DAGitty. Potential confounders are illustrated in pink. Potential mediating pathways are illustrated in green. Loneliness can lead to changes in health behaviors and increased risk of chronic disease.^1–5^ These factors may therefore be mediators on the causal pathway between loneliness and mortality (Figure 2b. A). However, health behaviors and health conditions have both been causally associated with loneliness and mortality (Figure 2b. B). Smoking could lead to feelings of loneliness through the neuro-pharmacological effects of nicotine.^1^ Alcohol and food culture often involves sharing experiences and conversation with members of one’s social network, which could minimize loneliness.^4,6^ Physical activity may reduce isolation by providing opportunities for social connection during activities, generating a sense of self-esteem, and improving mood.^7^ Poor sleep can lead to difficulty processing emotions and impact social relationships.^8^ Finaly, chronic conditions may reduce feelings of self-efficacy, limit participation in social activities and impact the quality of social relationships.^9,10^

## Appendix Text 1. Adjusted model specifications and covariate coding

**Model 0** was unadjusted.

**Model 1 adjusted** for age (continuous), biological sex (female; male), ethnicity (white; non-white), Townsend deprivation index (continuous), annual household income (<£18,000; £18,000 – £30,999; £31,000 – £51,999; >£52,000), population density (urban – population of 10,000 or more; non-urban – population of less than 10,000), commute mode of transportation (not employed; car-only; mixed transport including a car; mode excludes car), and month of assessment. Loneliness models were additionally adjusted for household size (living alone vs. not alone).

**Model 2 adjusted** for Model 1 covariates in addition to smoking status (current; previous; never).

**Model 3 adjusted** for Model 2 covariates in addition to alcohol consumption (frequent; special occasions; never), self-reported moderate-to-vigorous physical activity (MVPA) on a typical day (continuous, aggregated to minutes per week), sleep duration (continuous, in hours), red meat consumption (<2 times per week; 2-4 times per week; >2 times per week), processed meat consumption (<2 times per week; >2 times per week), oily fish consumption (<2 times per week; >2 times per week), non-oily fish consumption (<2 times per week; >2 times per week), cooked vegetable consumption (<2 servings per day; 2-4 servings per day; >4 servings per day), and fresh fruit consumption (<2 servings per day; 2-4 servings per day; >4 servings per day).

**Model 4** further adjusted for chronic conditions, including current depression (yes; no), long-standing illness, disability, or infirmity (yes; no), cancer (ever; never), waist circumference (continuous, measured in centimeters), and body mass index (BMI) (Normal; Obesity Class I; Obesity class II; Obesity class III; Overweight). The underweight BMI category had too few counts and was not considered in this analysis.

## Effect measure modifier: non-commute transportation mode

Participants reported on the most common transport modes used in the past four weeks (excluding commuting), with the option to select multiple modes. We categorized responses as:

1) Active transport only: if participants only selected walking and/or cycling
2) Public transport only: if participants only selected “public transport”
3) Mixed-without-car: if participants selected both active and public transportation modes, but not “car”
4) Mixed-with-car: if participants selected “car” in addition to an active and/or public transportation mode
5) Car-only: if participants only selected “car”

## Appendix Text 2. Methods used to generate additive interactions on an absolute risk scale

To improve interpretability of our main multiplicative interaction results, we estimated effect measure modification on the absolute risk scale using predictions derived from Cox proportional hazards models with interaction terms between social isolation and transportation mode, based on methods adapted from previous authors.^1^

We used the following method:

We fit Cox proportional hazards models for each mortality outcome, including main effects for social isolation and non-commute transportation mode, their interaction term, and adjusting for pre-specified covariates.

Using the fitted models, we predicted survival probabilities at 10 years for each joint exposure category. Predictions were generated from a standardized covariate profile, setting continuous covariates to their sample medians and categorical covariates to their sample modes.

Absolute 10-year risk of mortality was calculated using the following formula:^1^

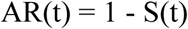

Where AR(t) denotes absolute risk of mortality at 10 years, and S(t) denotes predicted survival probability at 10 years.

Additive interaction was quantified using the interaction contrast (IC) on the absolute risk scale:^2^

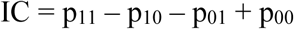

Where p_ij_ denotes the absolute risk for social isolation status i (0 = not isolated, 1 = isolated) and transportation mode j (0 = car-only, 1 = contrasting transportation mode)

Confidence intervals for interaction contrasts were obtained using model-based standard errors derived from the influence function (iid) output of the Cox model.

We multiplied ICs by 100,000 persons to estimate the number of excess deaths attributable to the additive interaction of social isolation and each transportation mode, beyond the sum of their individual effects.

Considerations for interpretation:

These additive interaction estimates were generated to improve the interpretability of our main multiplicative interaction results, rather than to serve as the primary test of interaction.

Confidence intervals were based on model-based standard errors. Resampling-based approaches, such as nonparametric bootstrapping, would provide more robust estimates of uncertainty.

Inference regarding statistical significance should therefore be interpreted with caution.

---

**Appendix Figure 4.**
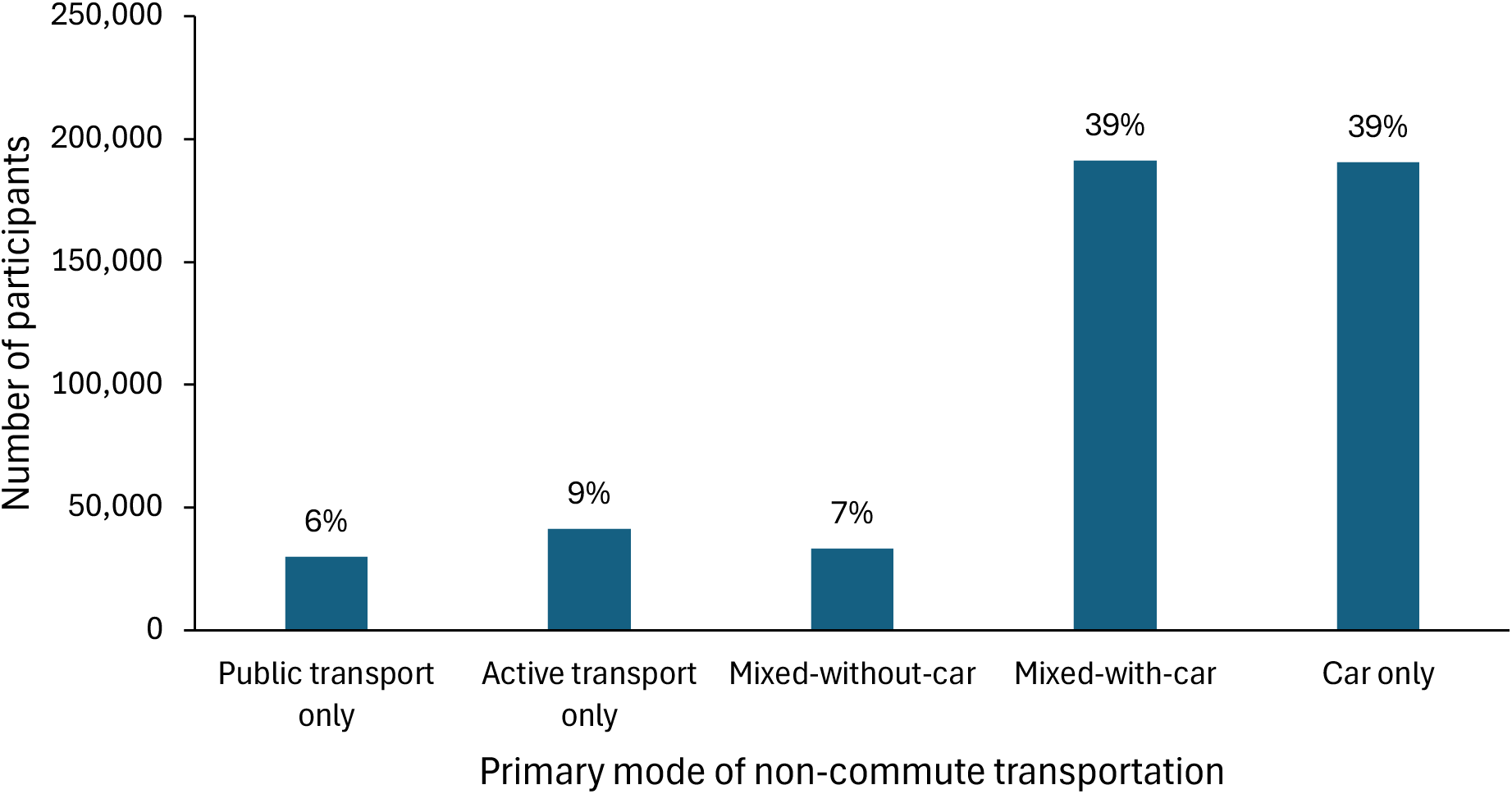
Primary mode of non-commute transportation in the UK Biobank

**Appendix Figure 5.**
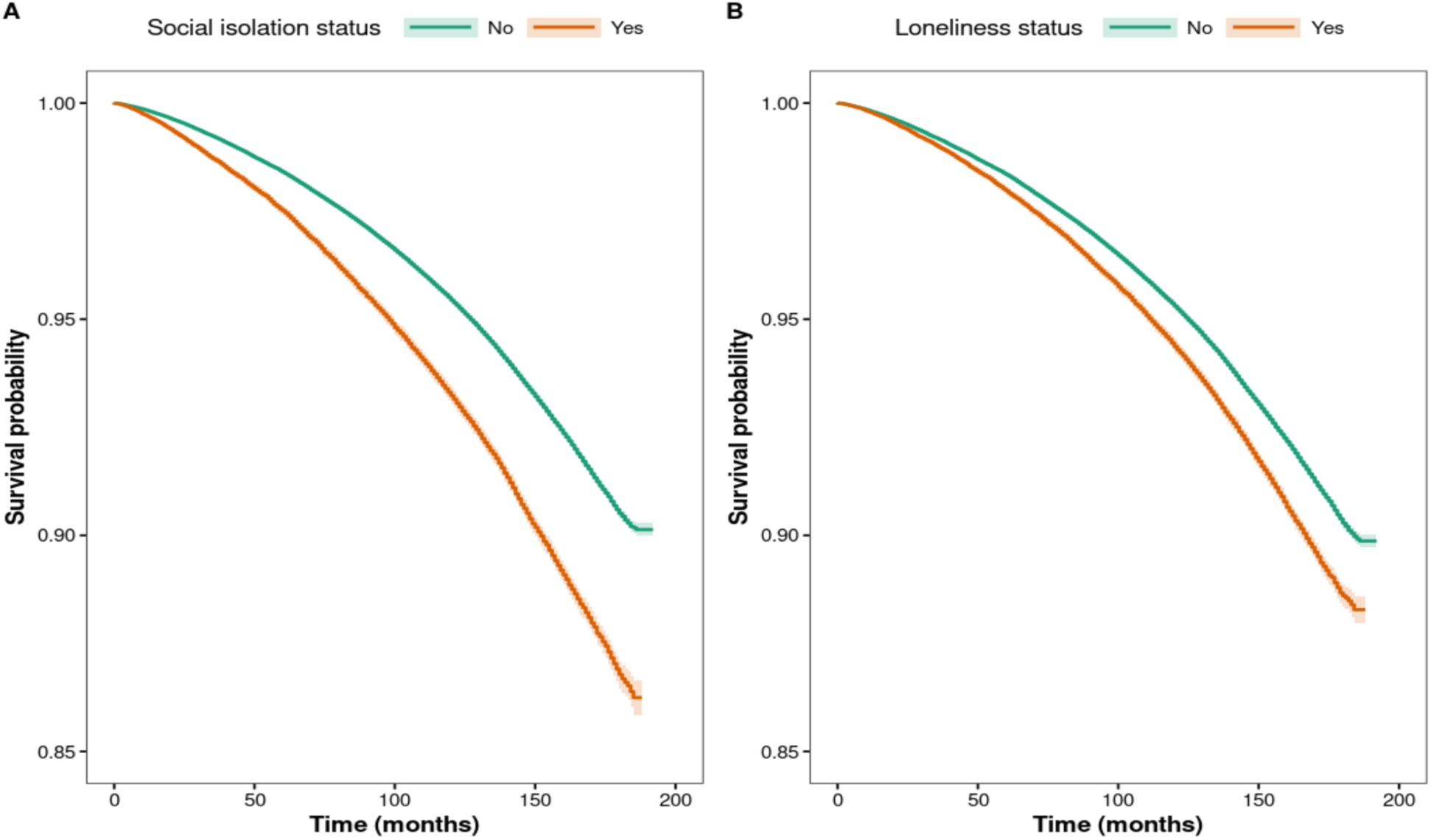
Kaplan-Meier curves of all-cause mortality by social isolation and loneliness status Kaplan-Meier curves of all-cause mortality by social isolation status. B) Kaplan-Meier curves of all-cause mortality by loneliness status.

**Appendix Figure 6.**
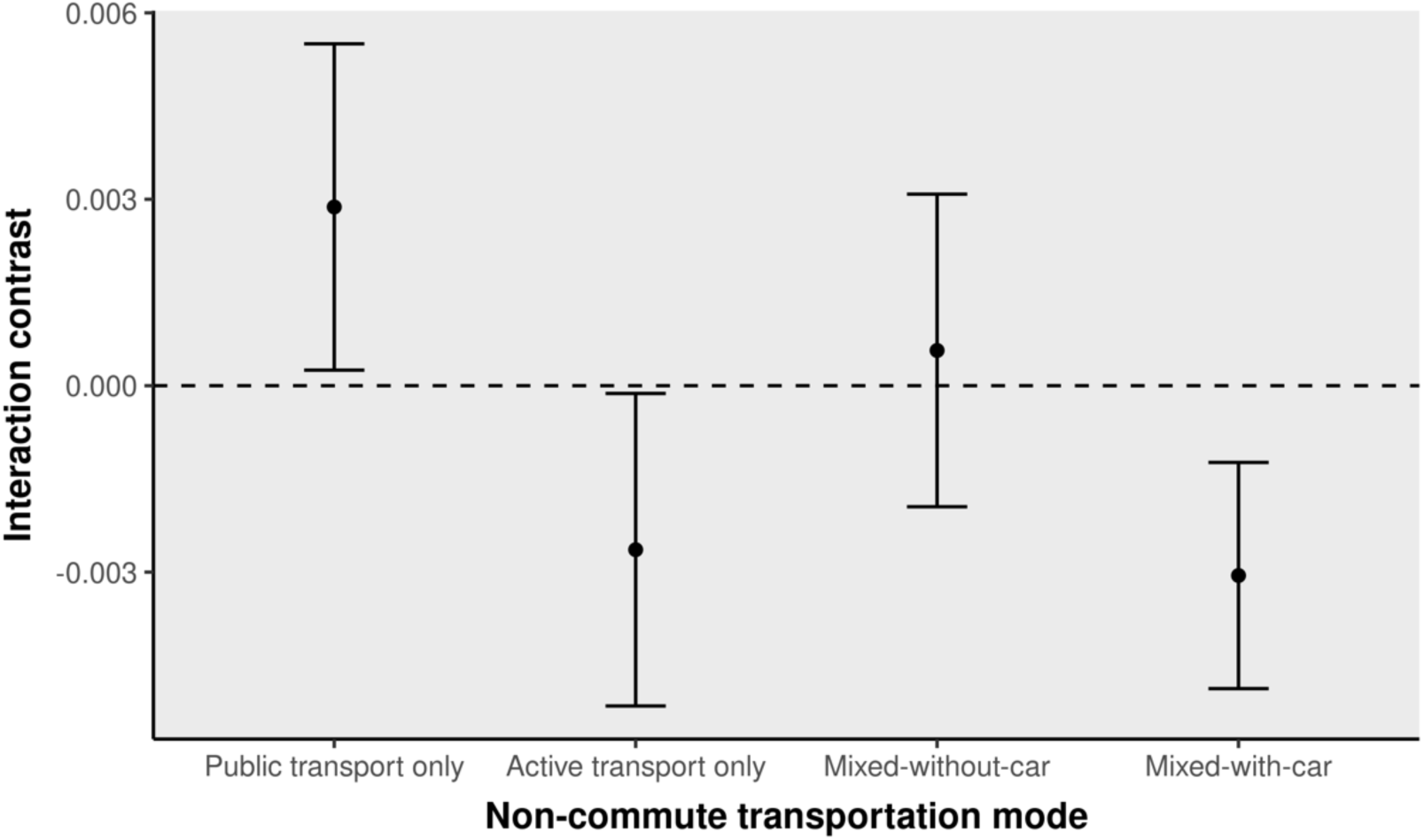
Interaction contrasts on the absolute risk scale between social isolation and non-commute transportation mode for 10-year all-cause mortality risk, using car-only transportation as the reference category

**Appendix Figure 7.**
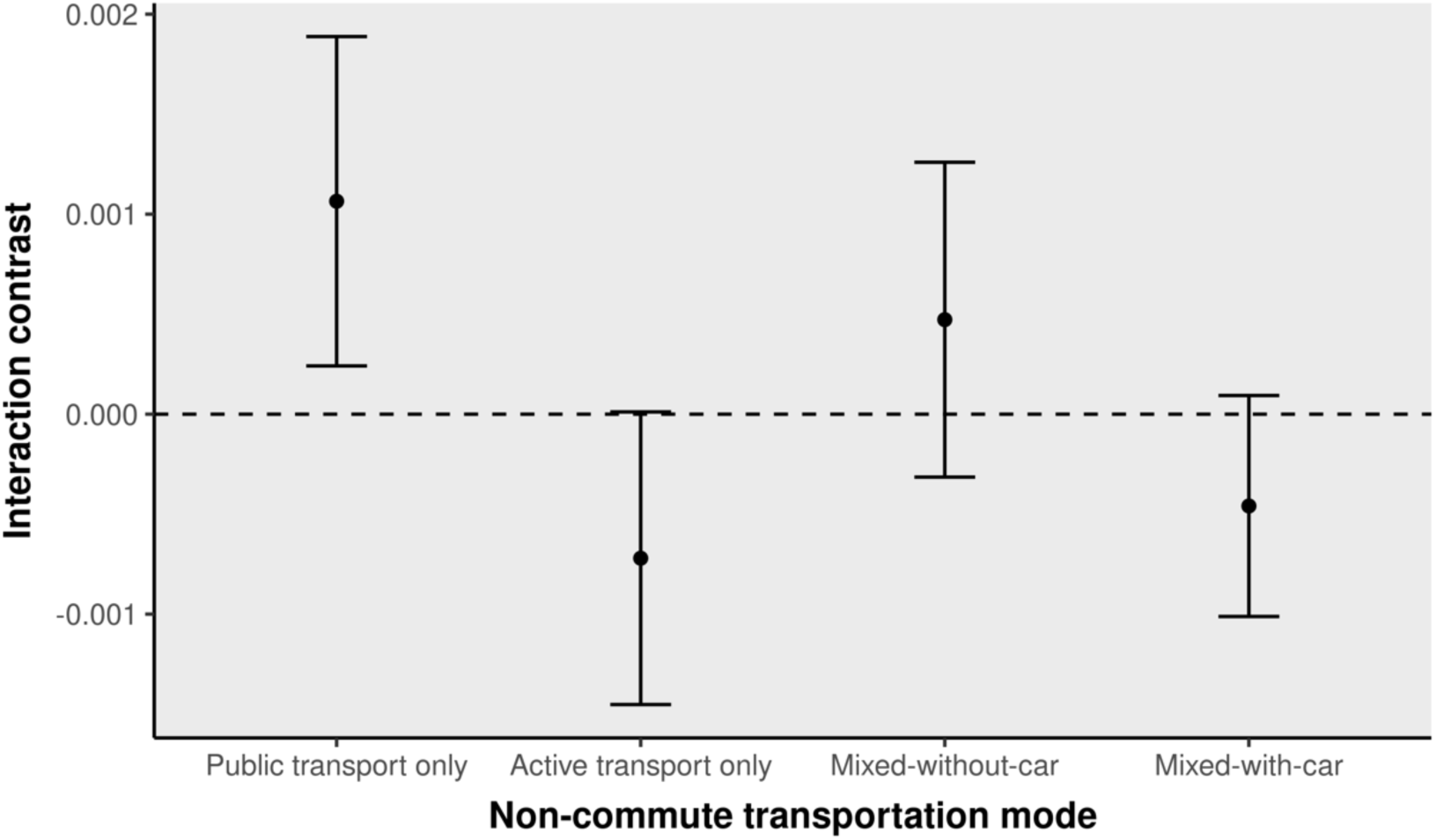
Interaction contrasts on the absolute risk scale between social isolation and non-commute transportation mode for 10-year cardiovascular disease mortality risk, using car-only transportation as the reference category

**Appendix Figure 8.**
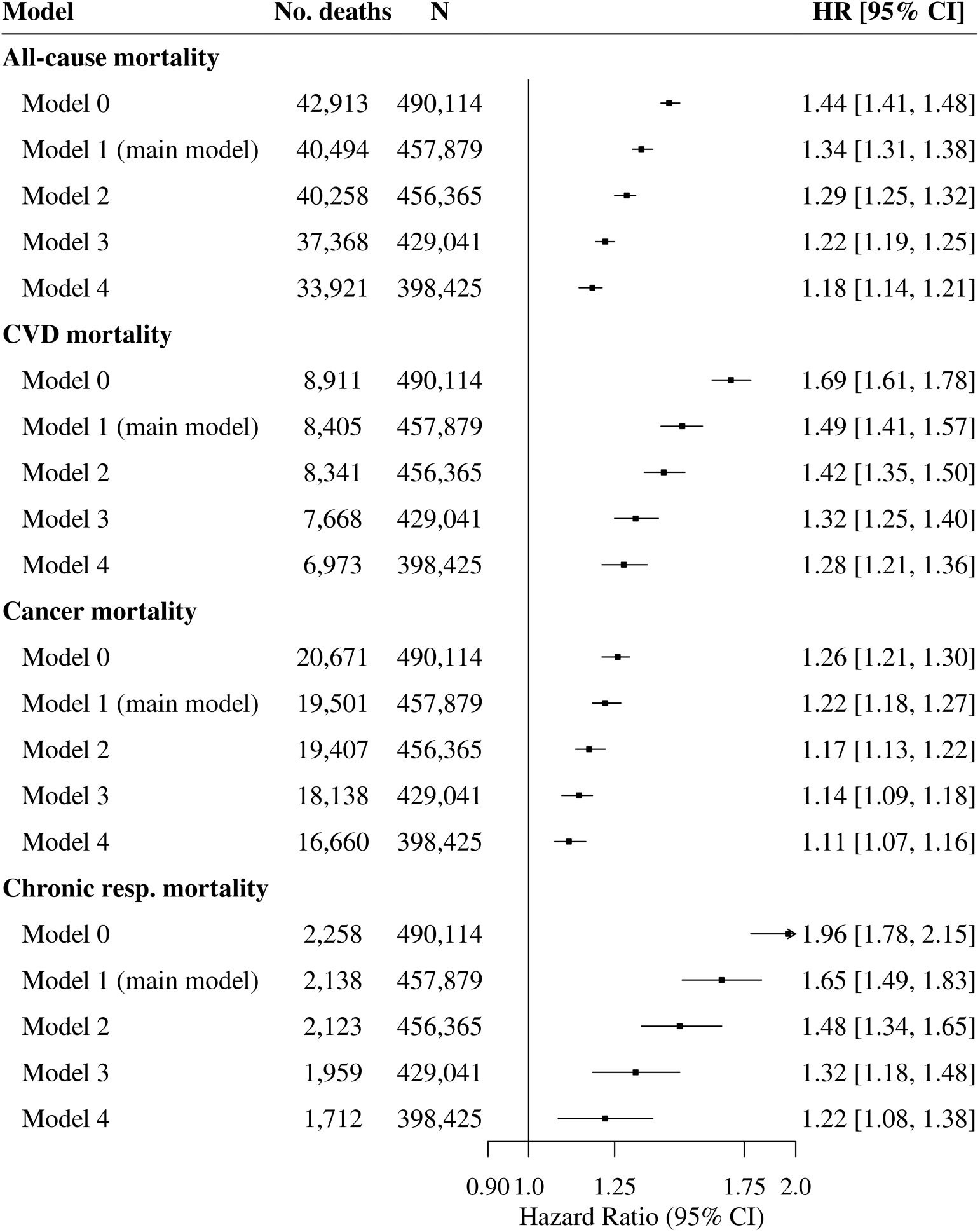
Hazard ratios and 35% confidence intervals for the association between social isolation and mortality outcomes across multiple adjusted models, without interactions between non-commute transportation mode and social isolation Model 0 unadjusted. Model 1 adjusted for age, sex, ethnicity, education, Townsend deprivation index, annual household income, month of assessment, commute transportation mode, and population density. Model 1 was used as the main model in analysis. Model 2 adjusted for Model 1 covariates in addition to smoking. Model 3 adjusted for Model 2 covariates in addition to alcohol consumption, physical activity, sleep, transportation, and diet. Model 4 adjusted for Model 3 covariates in addition to depression, long-standing illness, disability, or infirmity, cancer, waist circumference, and BMI. Abbreviations: No. deaths = Number of deaths; N = Total number of participants; HR = Hazard ratio; CI = Confidence interval; Ref. = Reference category. Arrowheads indicate confidence intervals extending beyond the displayed x-axis range.

**Appendix Table 2.** Association of social isolation with all-cause mortality by non-commute transport subgroup, using car-only as the reference category.

| Model | Model 1<sup>1</sup> |  | Model 2<sup>2</sup> |  | Model 3<sup>3</sup> |  | Model 4<sup>4</sup> |  |
| --- | --- | --- | --- | --- | --- | --- | --- | --- |
| N(deaths/total participants) | 39,766/455,072 |  | 39,539/453,592 |  | 36,702/426,540 |  | 33,456/396,466 |  |
|  | HR | 95% CI | HR | 95% CI | HR | 95% CI | HR | 95% CI |
| **Social isolation** |  |  |  |  |  |  |  |  |
| No | Ref. | - | Ref. | - | Ref. | - | Ref. | - |
| Yes | 1.34 | 1.28, 1.40 | 1.29 | 1.23, 1.35 | 1.22 | 1.17, 1.28 | 1.18 | 1.12, 1.23 |
| **Social isolation x transportation** |  |  |  |  |  |  |  |  |
| Yes x Public transport only | 1.05 | 0.97, 1.14 | 1.02 | 0.94, 1.10 | 1.02 | 0.94, 1.11 | 1.02 | 0.93, 1.11 |
| Yes x Active transport only | 0.94 | 0.86, 1.03 | 0.94 | 0.85, 1.03 | 0.94 | 0.85, 1.04 | 0.97 | 0.87, 1.07 |
| Yes x Mixed-without-car | 1.03 | 0.95, 1.12 | 1.04 | 0.96, 1.14 | 1.06 | 0.97, 1.15 | 1.07 | 0.97, 1.17 |
| Yes x Mixed-with-car | **0.93** | **0.87, 0.99** | **0.93** | **0.88, 1.00** | 0.95 | 0.88, 1.01 | 0.95 | 0.89, 1.02 |
| **Transportation mode** |  |  |  |  |  |  |  |  |
| Car-only | Ref. | - | Ref. | - | Ref. | - | Ref. | - |
| Public transport only | 1.13 | 1.08, 1.18 | 1.13 | 1.09, 1.18 | 1.11 | 1.06, 1.17 | 1.13 | 1.08, 1.19 |
| Active transport only | 0.88 | 0.84, 0.92 | 0.89 | 0.85, 0.93 | 0.92 | 0.88, 0.96 | 1.01 | 0.96, 1.06 |
| Mixed-without-car | 0.95 | 0.91, 0.99 | 0.96 | 0.92, 1.01 | 0.98 | 0.93, 1.02 | 1.03 | 0.98, 1.08 |
| Mixed-with-car | 0.86 | 0.84, 0.88 | 0.88 | 0.86, 0.91 | 0.91 | 0.89, 0.94 | 0.96 | 0.93, 0.98 |

Model 1 adjusted for age, sex, ethnicity, education, Townsend deprivation index, annual household income, month of assessment, commute transportation mode, and population density. Model 1 was used as the main model in analysis.
Model 2 adjusted for Model 1 covariates in addition to smoking.
Model 3 adjusted for Model 2 covariates in addition to alcohol consumption, physical activity, sleep, transportation, and diet.
Model 4 adjusted for Model 3 covariates in addition to depression, long-standing illness, disability, or infirmity, cancer, waist circumference, and BMI.
Abbreviations: N = Number of participants; HR = Hazard ratio; CI = Confidence interval; Ref. = Reference category

**Appendix Table 3.** Association of social isolation with cardiovascular disease mortality by non-commute transport subgroup, using car-only as the reference category.

| Model<br>N(deaths/total participants) | Model 1<sup>1</sup> |  | Model 2<sup>2</sup> |  | Model 3<sup>3</sup> |  | Model 4<sup>4</sup> |  |
| --- | --- | --- | --- | --- | --- | --- | --- | --- |
|  | 8,232/455,072 |  | 8,169/453,592 |  | 7,512/426,540 |  | 6,855/396,466 |  |
|  | HR | 95% CI | HR | 95% CI | HR | 95% CI | HR | 95% CI |
| **Social isolation** |  |  |  |  |  |  |  |  |
| No | Ref. | - | Ref. | - | Ref. | - | Ref. | - |
| Yes | 1.44 | 1.32, 1.58 | 1.39 | 1.27, 1.53 | 1.31 | 1.19, 1.44 | 1.27 | 1.15, 1.40 |
| **Social isolation x transportation** |  |  |  |  |  |  |  |  |
| Yes x Public transport only | **1.17** | **1.00, 1.37** | 1.12 | 0.95, 1.32 | 1.11 | 0.94, 1.32 | 1.12 | 0.93, 1.34 |
| Yes x Active transport only | 0.88 | 0.71, 1.08 | 0.86 | 0.70, 1.05 | 0.84 | 0.67, 1.05 | 0.84 | 0.66, 1.06 |
| Yes x Mixed-without-car | 1.11 | 0.93, 1.32 | 1.11 | 0.93, 1.32 | 1.12 | 0.93, 1.34 | 1.11 | 0.91, 1.35 |
| Yes x Mixed-with-car | 0.94 | 0.82, 1.08 | 0.94 | 0.82, 1.08 | 0.93 | 0.80, 1.07 | 0.95 | 0.82, 1.11 |
| **Transportation mode** |  |  |  |  |  |  |  |  |
| Car-only | Ref. | - | Ref. | - | Ref. | - | Ref. | - |
| Public transport only | 1.13 | 1.02, 1.24 | 1.14 | 1.03, 1.25 | 1.12 | 1.01, 1.24 | 1.13 | 1.02, 1.26 |
| Active transport only | 0.83 | 0.75, 0.92 | 0.85 | 0.77, 0.94 | 0.86 | 0.77, 0.96 | 1.00 | 0.89, 1.12 |
| Mixed-without-car | 0.98 | 0.88, 1.08 | 0.99 | 0.90, 1.10 | 1.01 | 0.91, 1.12 | 1.10 | 0.99, 1.23 |
| Mixed-with-car | 0.82 | 0.78, 0.87 | 0.85 | 0.80, 0.90 | 0.88 | 0.83, 0.93 | 0.94 | 0.88, 1.00 |

Model 1 adjusted for age, sex, ethnicity, education, Townsend deprivation index, annual household income, month of assessment, commute transportation mode, and population density. Model 1 was used as the main model in analysis.
Model 2 adjusted for Model 1 covariates in addition to smoking.
Model 3 adjusted for Model 2 covariates in addition to alcohol consumption, physical activity, sleep, transportation, and diet.
Model 4 adjusted for Model 3 covariates in addition to depression, long-standing illness, disability, or infirmity, cancer, waist circumference, and BMI.
Abbreviations: N = Number of participants; HR = Hazard ratio; CI = Confidence interval; Ref. = Reference category

**Appendix Table 4.** Association of social isolation with cancer mortality by non-commute transport subgroup, using car-only as the reference category.

| Model | Model 1<sup>1</sup> |  | Model 2<sup>2</sup> |  | Model 3<sup>3</sup> |  | Model 4<sup>4</sup> |  |
| --- | --- | --- | --- | --- | --- | --- | --- | --- |
| N(deaths/total participants) | 19,317/455,072 |  | 19,224/453,592 |  | 17,966/426,540 |  | 16,529/396,466 |  |
|  | HR | 95% CI | HR | 95% CI | HR | 95% CI | HR | 95% CI |
| **Social isolation** |  |  |  |  |  |  |  |  |
| No | Ref. | - | Ref. | - | Ref. | - | Ref. | - |
| Yes | 1.23 | 1.15, 1.31 | 1.18 | 1.11, 1.26 | 1.15 | 1.08, 1.23 | 1.12 | 1.05, 1.20 |
| **Social isolation x transportation** |  |  |  |  |  |  |  |  |
| Yes x Public transport only | 1.01 | 0.89, 1.14 | 0.99 | 0.87, 1.12 | 0.98 | 0.86, 1.12 | 0.96 | 0.83, 1.10 |
| Yes x Active transport only | 0.95 | 0.83, 1.09 | 0.95 | 0.83, 1.09 | 0.92 | 0.79, 1.06 | 0.96 | 0.82, 1.12 |
| Yes x Mixed-without-car | 1.02 | 0.90, 1.16 | 1.04 | 0.91, 1.18 | 1.03 | 0.90, 1.17 | 1.04 | 0.90, 1.20 |
| Yes x Mixed-with-car | 0.96 | 0.87, 1.05 | 0.97 | 0.88, 1.06 | 0.97 | 0.88, 1.07 | 0.98 | 0.89, 1.09 |
| **Transportation mode** |  |  |  |  |  |  |  |  |
| Car-only | Ref. | - | Ref. | - | Ref. | - | Ref. | - |
| Public transport only | 1.04 | 0.98, 1.11 | 1.04 | 0.98, 1.12 | 1.04 | 0.97, 1.11 | 1.07 | 0.99, 1.15 |
| Active transport only | 0.89 | 0.84, 0.95 | 0.91 | 0.85, 0.97 | 0.94 | 0.88, 1.01 | 1.01 | 0.95, 1.09 |
| Mixed-without-car | 0.94 | 0.88, 1.00 | 0.95 | 0.89, 1.01 | 0.95 | 0.89, 1.02 | 0.99 | 0.92, 1.07 |
| Mixed-with-car | 0.92 | 0.89, 0.96 | 0.95 | 0.91, 0.98 | 0.97 | 0.93, 1.00 | 0.99 | 0.96, 1.03 |

Model 3 adjusted for Model 2 covariates in addition to alcohol consumption, physical activity, sleep, transportation, and diet.
Model 4 adjusted for Model 3 covariates in addition to depression, long-standing illness, disability, or infirmity, cancer, waist circumference, and BMI.
Abbreviations: N = Number of participants; HR = Hazard ratio; CI = Confidence interval; Ref. = Reference category

**Appendix Table 5.** Association of social isolation with chronic respiratory mortality by non-commute transport subgroup, using car-only as the reference category.

| Model<br>N(deaths/total participants) | Model 1<sup>1</sup><br>2,072/455,072 |  | Model 2<sup>2</sup><br>2,060/453,592 |  | Model 3<sup>3</sup><br>1,901/426,540 |  | Model 4<sup>4</sup><br>1,672/396,466 |  |
| --- | --- | --- | --- | --- | --- | --- | --- | --- |
|  | HR | 95% CI | HR | 95% CI | HR | 95% CI | HR | 95% CI |
| **Social isolation** |  |  |  |  |  |  |  |  |
| No | Ref. | - | Ref. | - | Ref. | - | Ref. | - |
| Yes | 1.65 | 1.39, 1.95 | 1.49 | 1.26, 1.76 | 1.29 | 1.07, 1.54 | 1.16 | 0.95, 1.41 |
| **Social isolation x transportation** |  |  |  |  |  |  |  |  |
| Yes x Public transport only | 1.00 | 0.76, 1.32 | 0.96 | 0.73, 1.27 | 0.98 | 0.73, 1.32 | 1.09 | 0.79, 1.51 |
| Yes x Active transport only | 1.17 | 0.76, 1.79 | 1.15 | 0.75, 1.78 | 1.31 | 0.83, 2.06 | 1.31 | 0.80, 2.17 |
| Yes x Mixed-without-car | 0.88 | 0.62, 1.24 | 0.93 | 0.66, 1.32 | 0.99 | 0.69, 1.42 | 1.03 | 0.69, 1.53 |
| Yes x Mixed-with-car | 0.98 | 0.75, 1.28 | 0.99 | 0.76, 1.30 | 1.08 | 0.81, 1.43 | 1.12 | 0.83, 1.52 |
| **Transportation mode** |  |  |  |  |  |  |  |  |
| Car-only | Ref. | - | Ref. | - | Ref. | - | Ref. | - |
| Public transport only | 1.12 | 0.95, 1.33 | 1.13 | 0.95, 1.33 | 1.10 | 0.93, 1.32 | 1.07 | 0.88, 1.29 |
| Active transport only | 0.47 | 0.36, 0.59 | 0.48 | 0.38, 0.62 | 0.50 | 0.38, 0.65 | 0.53 | 0.40, 0.71 |
| Mixed-without-car | 0.78 | 0.64, 0.94 | 0.78 | 0.64, 0.95 | 0.81 | 0.66, 1.00 | 0.83 | 0.67, 1.04 |
| Mixed-with-car | 0.63 | 0.56, 0.71 | 0.67 | 0.60, 0.76 | 0.69 | 0.61, 0.78 | 0.72 | 0.64, 0.82 |

Abbreviations: N = Number of participants; HR = Hazard ratio; CI = Confidence interval; Ref. = Reference category

**Appendix Table 6.** Estimated 10-year excess deaths per 100,000 persons attributable to the additive interaction between social isolation and non-commute transportation modes.

| Non-commute<br>transportation mode | All-cause mortality |  | Cardiovascular disease<br>mortality |  |
| --- | --- | --- | --- | --- |
|  | Excess deaths<br>per 100,000 | Additive<br>interaction<br>type | Excess deaths<br>per 100,000 | Additive<br>interaction<br>type |
| Car-only (reference) |  |  |  |  |
| Public transport only | 288 | Super-additive | 106 | Super-additive |
| Active transport only | -264 | Sub-additive | -72 | Sub-additive |
| Mixed-without-car | 57 | Super-additive | 47 | Super-additive |
| Mixed-with-car | -305 | Sub-additive | -46 | Sub-additive |

**Appendix Figure 9.**
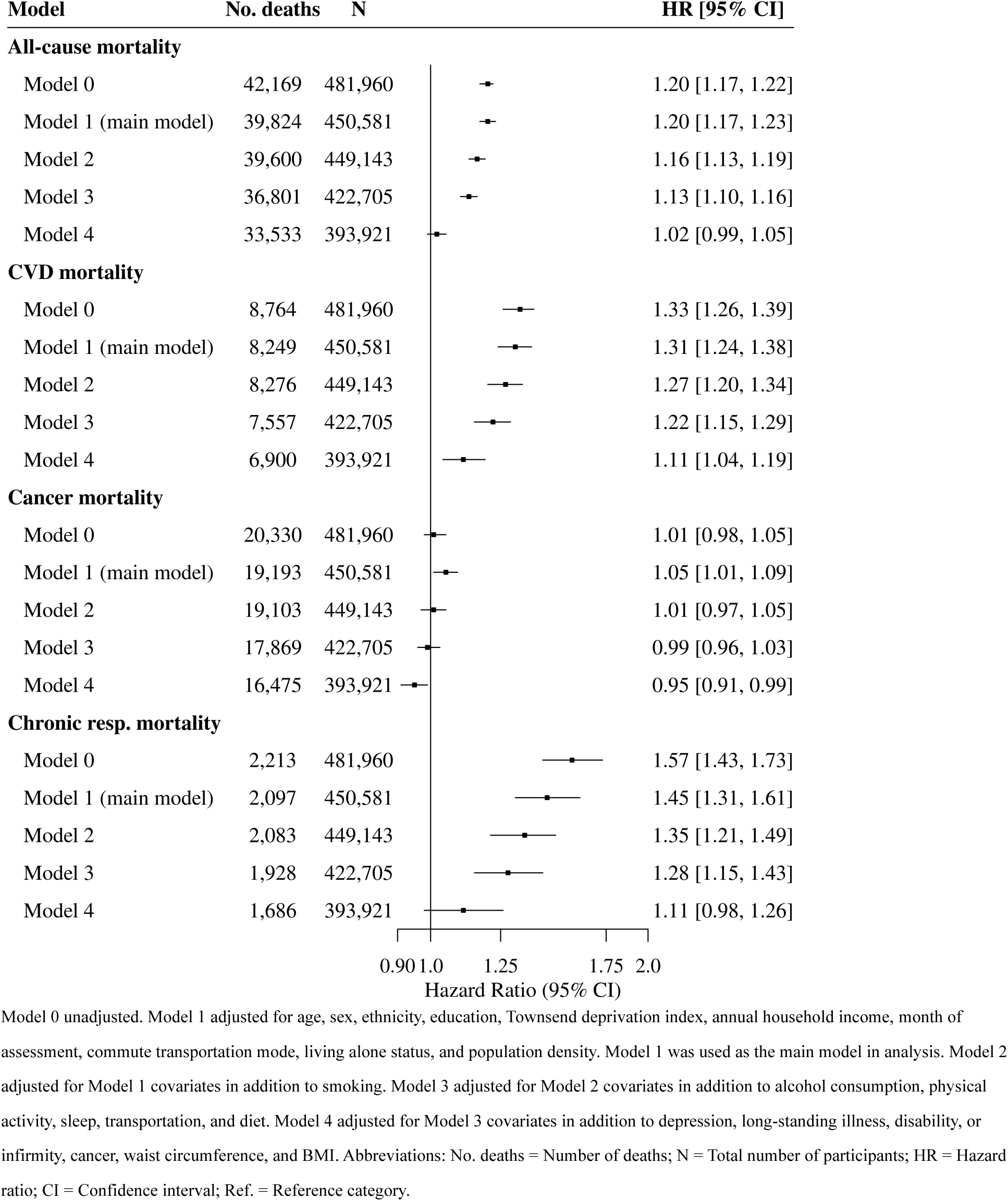
Hazard ratios and 35% confidence intervals for the association between loneliness and mortality outcomes across multiple adjusted models, without interactions between non-commute transportation mode and loneliness Model 0 unadjusted. Model 1 adjusted for age, sex, ethnicity, education, Townsend deprivation index, annual household income, month of assessment, commute transportation mode, living alone status, and population density. Model 1 was used as the main model in analysis. Model 2 adjusted for Model 1 covariates in addition to smoking. Model 3 adjusted for Model 2 covariates in addition to alcohol consumption, physical activity, sleep, transportation, and diet. Model 4 adjusted for Model 3 covariates in addition to depression, long-standing illness, disability, or infirmity, cancer, waist circumference, and BMI. Abbreviations: No. deaths = Number of deaths; N = Total number of participants; HR = Hazard ratio; CI = Confidence interval; Ref. = Reference category.

**Appendix Table 7.** Association of loneliness with all-cause mortality by non-commute transport subgroup, using car-only as the reference category.

| Model | Model 1<sup>1</sup> |  | Model 2<sup>2</sup> |  | Model 3<sup>3</sup> |  | Model 4<sup>4</sup> |  |
| --- | --- | --- | --- | --- | --- | --- | --- | --- |
| N(deaths/total participants) | 39,113/447,862 |  | 38,898/446,454 |  | 36,148/420,263 |  | 33,077/392,004 |  |
|  | HR | 95% CI | HR | 95% CI | HR | 95% CI | HR | 95% CI |
| **Loneliness** |  |  |  |  |  |  |  |  |
| No | Ref. | - | Ref. | - | Ref. | - | Ref. | - |
| Yes | 1.19 | 1.14, 1.24 | 1.15 | 1.10, 1.20 | 1.13 | 1.08, 1.18 | 1.02 | 0.97, 1.07 |
| **Loneliness x transportation** |  |  |  |  |  |  |  |  |
| Yes x Public transport only | 0.97 | 0.90, 1.05 | 0.96 | 0.89, 1.04 | 0.95 | 0.88, 1.03 | 0.94 | 0.86, 1.02 |
| Yes x Active transport only | 0.98 | 0.89, 1.08 | 0.96 | 0.87, 1.06 | 0.96 | 0.87, 1.07 | 1.00 | 0.90, 1.12 |
| Yes x Mixed-without-car | 1.03 | 0.95, 1.13 | 1.03 | 0.94, 1.12 | 1.02 | 0.93, 1.12 | 1.05 | 0.96, 1.16 |
| Yes x Mixed-with-car | 0.97 | 0.92, 1.03 | 0.99 | 0.93, 1.05 | 0.98 | 0.92, 1.04 | 0.98 | 0.92, 1.05 |
| **Transportation mode** |  |  |  |  |  |  |  |  |
| Car-only | Ref. | - | Ref. | - | Ref. | - | Ref. | - |
| Public transport only | 1.15 | 1.10, 1.20 | 1.15 | 1.10, 1.20 | 1.13 | 1.08, 1.18 | 1.15 | 1.10, 1.21 |
| Active transport only | 0.87 | 0.83, 0.91 | 0.88 | 0.85, 0.93 | 0.91 | 0.87, 0.96 | 1.00 | 0.95, 1.05 |
| Mixed-without-car | 0.94 | 0.90, 0.99 | 0.96 | 0.92, 1.00 | 0.98 | 0.93, 1.03 | 1.02 | 0.97, 1.08 |
| Mixed-with-car | 0.86 | 0.83, 0.88 | 0.88 | 0.85, 0.90 | 0.91 | 0.88, 0.93 | 0.95 | 0.92, 0.98 |

Model 1 adjusted for age, sex, ethnicity, education, Townsend deprivation index, annual household income, month of assessment, commute transportation mode, living alone status, and population density. Model 1 was used as the main model in analysis.
Model 2 adjusted for Model 1 covariates in addition to smoking.
Model 3 adjusted for Model 2 covariates in addition to alcohol consumption, physical activity, sleep, transportation, and diet.
Model 4 adjusted for Model 3 covariates in addition to depression, long-standing illness, disability, or infirmity, cancer, waist circumference, and BMI.
Abbreviations: N = Number of participants; HR = Hazard ratio; CI = Confidence interval; Ref. = Reference category

**Appendix Table 8.** Association of loneliness with cardiovascular disease mortality by non-commute transport subgroup, using car-only as the reference category.

| Model<br>N(deaths/total participants) | Model 1<sup>1</sup><br>8,108/447,862 |  | Model 2<sup>2</sup><br>8,048/446,454 |  | Model 3<sup>3</sup><br>7,405/420,263 |  | Model 4<sup>4</sup><br>6,785/392,004 |  |
| --- | --- | --- | --- | --- | --- | --- | --- | --- |
|  | HR | 95% CI | HR | 95% CI | HR | 95% CI | HR | 95% CI |
| **Loneliness** |  |  |  |  |  |  |  |  |
| No | Ref. | - | Ref. | - | Ref. | - | Ref. | - |
| Yes | 1.22 | 1.12, 1.33 | 1.19 | 1.08, 1.30 | 1.15 | 1.05, 1.26 | 1.06 | 0.96, 1.17 |
| **Loneliness x transportation** |  |  |  |  |  |  |  |  |
| Yes x Public transport only | 1.05 | 0.90, 1.24 | 1.04 | 0.88, 1.22 | 1.00 | 0.85, 1.19 | 0.97 | 0.81, 1.16 |
| Yes x Active transport only | **1.23** | **1.00, 1.51** | 1.2 | 0.98, 1.48 | 1.21 | 0.97, 1.51 | **1.28** | **1.01, 1.61** |
| Yes x Mixed-without-car | 1.16 | 0.97, 1.39 | 1.15 | 0.96, 1.38 | 1.17 | 0.97, 1.41 | **1.22** | **1.00, 1.48** |
| Yes x Mixed-with-car | 1.03 | 0.90, 1.17 | 1.04 | 0.92, 1.19 | 1.04 | 0.91, 1.19 | 1.02 | 0.89, 1.18 |
| **Transportation mode** |  |  |  |  |  |  |  |  |
| Car-only | Ref. | - | Ref. | - | Ref. | - | Ref. | - |
| Public transport only | 1.14 | 1.04, 1.26 | 1.15 | 1.04, 1.27 | 1.14 | 1.03, 1.26 | 1.17 | 1.05, 1.30 |
| Active transport only | 0.75 | 0.68, 0.84 | 0.78 | 0.70, 0.86 | 0.78 | 0.70, 0.87 | 0.89 | 0.79, 1.00 |
| Mixed-without-car | 0.95 | 0.86, 1.04 | 0.97 | 0.87, 1.07 | 0.98 | 0.88, 1.08 | 1.05 | 0.94, 1.18 |
| Mixed-with-car | 0.81 | 0.76, 0.86 | 0.83 | 0.78, 0.88 | 0.86 | 0.81, 0.91 | 0.92 | 0.86, 0.98 |

Model 1 adjusted for age, sex, ethnicity, education, Townsend deprivation index, annual household income, month of assessment, commute transportation mode, living alone status, and population density. Model 1 was used as the main model in analysis.
Model 2 adjusted for Model 1 covariates in addition to smoking.
Model 3 adjusted for Model 2 covariates in addition to alcohol consumption, physical activity, sleep, transportation, and diet.
Model 4 adjusted for Model 3 covariates in addition to depression, long-standing illness, disability, or infirmity, cancer, waist circumference, and BMI.
Abbreviations: N = Number of participants; HR = Hazard ratio; CI = Confidence interval; Ref. = Reference category

**Appendix Table 9.** Association of loneliness with cancer disease mortality by non-commute transport subgroup, using car-only as the reference category.

| Model | Model 1<sup>1</sup> |  | Model 2<sup>2</sup> |  | Model 3<sup>3</sup> |  | Model 4<sup>4</sup> |  |
| --- | --- | --- | --- | --- | --- | --- | --- | --- |
| N(deaths/total participants) | 19,014/447,862 |  | 18,925/446,454 |  | 17,702/420,263 |  | 16,348/392,004 |  |
|  | HR | 95% CI | HR | 95% CI | HR | 95% CI | HR | 95% CI |
| **Loneliness** |  |  |  |  |  |  |  |  |
| No | Ref. | - | Ref. | - | Ref. | - | Ref. | - |
| Yes | 1.05 | 0.99, 1.12 | 1.01 | 0.95, 1.08 | 1.01 | 0.95, 1.08 | 0.96 | 0.90, 1.03 |
| **Loneliness x transportation** |  |  |  |  |  |  |  |  |
| Yes x Public transport only | 0.95 | 0.84, 1.08 | 0.94 | 0.83, 1.06 | 0.94 | 0.82, 1.07 | 0.92 | 0.80, 1.06 |
| Yes x Active transport only | 0.99 | 0.85, 1.14 | 0.97 | 0.84, 1.12 | 0.97 | 0.83, 1.13 | 1.00 | 0.85, 1.17 |
| Yes x Mixed-without-car | 0.91 | 0.80, 1.05 | 0.91 | 0.80, 1.04 | 0.87 | 0.76, 1.01 | 0.89 | 0.77, 1.04 |
| Yes x Mixed-with-car | 1.01 | 0.93, 1.10 | 1.03 | 0.94, 1.12 | 1.01 | 0.93, 1.11 | 1.00 | 0.91, 1.10 |
| **Transportation mode** |  |  |  |  |  |  |  |  |
| Car-only | Ref. | - | Ref. | - | Ref. | - | Ref. | - |
| Public transport only | 1.06 | 0.99, 1.14 | 1.06 | 1.00, 1.14 | 1.06 | 0.99, 1.13 | 1.08 | 1.00, 1.16 |
| Active transport only | 0.89 | 0.83, 0.95 | 0.91 | 0.85, 0.96 | 0.93 | 0.87, 1.00 | 1.01 | 0.94, 1.08 |
| Mixed-without-car | 0.96 | 0.90, 1.03 | 0.98 | 0.92, 1.05 | 0.99 | 0.93, 1.06 | 1.03 | 0.96, 1.10 |
| Mixed-with-car | 0.92 | 0.88, 0.95 | 0.94 | 0.90, 0.97 | 0.96 | 0.93, 1.00 | 0.99 | 0.95, 1.03 |

Model 1 adjusted for age, sex, ethnicity, education, Townsend deprivation index, annual household income, month of assessment, commute transportation mode, living alone status, and population density. Model 1 was used as the main model in analysis.
Model 2 adjusted for Model 1 covariates in addition to smoking.
Model 3 adjusted for Model 2 covariates in addition to alcohol consumption, physical activity, sleep, transportation, and diet.
Model 4 adjusted for Model 3 covariates in addition to depression, long-standing illness, disability, or infirmity, cancer, waist circumference, and BMI.
Abbreviations: N = Number of participants; HR = Hazard ratio; CI = Confidence interval; Ref. = Reference category

**Appendix Table 10.** Association of loneliness with chronic respiratory mortality by non-commute transport subgroup, using car-only as the reference category.

| Model<br>N(deaths/total participants) | Model 1<sup>1</sup><br>2,033/447,862 |  | Model 2<sup>2</sup><br>2,022/446,454 |  | Model 3<sup>3</sup><br>1,871/420,263 |  | Model 4<sup>4</sup><br>1,647/392,004 |  |
| --- | --- | --- | --- | --- | --- | --- | --- | --- |
|  | HR | 95% CI | HR | 95% CI | HR | 95% CI | HR | 95% CI |
| **Loneliness** |  |  |  |  |  |  |  |  |
| No | Ref. | - | Ref. | - | Ref. | - | Ref. | - |
| Yes | 1.46 | 1.25, 1.72 | 1.36 | 1.15, 1.59 | 1.31 | 1.11, 1.54 | 1.16 | 0.96, 1.40 |
| **Loneliness x transportation** |  |  |  |  |  |  |  |  |
| Yes x Public transport only | 0.95 | 0.72, 1.25 | 0.93 | 0.71, 1.23 | 0.91 | 0.68, 1.22 | 0.94 | 0.69, 1.29 |
| Yes x Active transport only | 0.72 | 0.44, 1.17 | 0.68 | 0.42, 1.12 | 0.62 | 0.36, 1.07 | 0.58 | 0.32, 1.06 |
| Yes x Mixed-without-car | 1.00 | 0.70, 1.41 | 1.01 | 0.72, 1.44 | 0.97 | 0.68, 1.40 | 0.91 | 0.61, 1.37 |
| Yes x Mixed-with-car | 0.99 | 0.77, 1.28 | 1.02 | 0.79, 1.32 | 1.04 | 0.80, 1.35 | 1.00 | 0.75, 1.32 |
| **Transportation mode** |  |  |  |  |  |  |  |  |
| Car-only | Ref. | - | Ref. | - | Ref. | - | Ref. | - |
| Public transport only | 1.14 | 0.96, 1.35 | 1.14 | 0.96, 1.35 | 1.12 | 0.94, 1.34 | 1.10 | 0.90, 1.33 |
| Active transport only | 0.54 | 0.43, 0.68 | 0.56 | 0.45, 0.71 | 0.61 | 0.48, 0.78 | 0.65 | 0.50, 0.85 |
| Mixed-without-car | 0.74 | 0.60, 0.90 | 0.75 | 0.61, 0.92 | 0.80 | 0.65, 0.98 | 0.83 | 0.67, 1.04 |
| Mixed-with-car | 0.62 | 0.55, 0.70 | 0.66 | 0.59, 0.75 | 0.68 | 0.60, 0.78 | 0.73 | 0.64, 0.83 |

Model 1 adjusted for age, sex, ethnicity, education, Townsend deprivation index, annual household income, month of assessment, commute transportation mode, living alone status, and population density. Model 1 was used as the main model in analysis.
Model 2 adjusted for Model 1 covariates in addition to smoking.
Model 3 adjusted for Model 2 covariates in addition to alcohol consumption, physical activity, sleep, transportation, and diet.
Model 4 adjusted for Model 3 covariates in addition to depression, long-standing illness, disability, or infirmity, cancer, waist circumference, and BMI.
Abbreviations: N = Number of participants; HR = Hazard ratio; CI = Confidence interval; Ref. = Reference category

**Appendix Table 11.** Association of social isolation with all-cause and cause-specific mortality by non-commute transport subgroup, using car-only as the reference category and not adjusted for commuting transportation mode.

| Mortality outcome<br>N(deaths/total participants) | All-cause<br>41,080/475,842 |  | CVD<br>8,487/475,842 |  | Cancer<br>20,013/475,842 |  | Chronic respiratory<br>2,122/475,842 |  |
| --- | --- | --- | --- | --- | --- | --- | --- | --- |
|  | HR | 95% CI | HR | 95% CI | HR | 95% CI | HR | 95% CI |
| **Social isolation** |  |  |  |  |  |  |  |  |
| No | Ref. | - | Ref. | - | Ref. | - | Ref. | - |
| Yes | 1.32 | 1.26, 1.38 | 1.43 | 1.31, 1.56 | 1.23 | 1.15, 1.31 | 1.61 | 1.36, 1.89 |
| **Social isolation x**<br>**transportation** |  |  |  |  |  |  |  |  |
| Yes x Public transport only | 1.06 | 0.99, 1.15 | **1.17** | **1.00, 1.37** | 1.01 | 0.89, 1.14 | 1.04 | 0.79, 1.37 |
| Yes x Active transport only | 0.96 | 0.88, 1.05 | 0.89 | 0.73, 1.09 | 0.97 | 0.85, 1.11 | 1.19 | 0.78, 1.82 |
| Yes x Mixed-without-car | 1.03 | 0.95, 1.12 | 1.12 | 0.94, 1.33 | 1.01 | 0.89, 1.15 | 0.88 | 0.62, 1.24 |
| Yes x Mixed-with-car | **0.93** | **0.87, 0.99** | 0.92 | 0.81, 1.06 | 0.96 | 0.87, 1.05 | 0.93 | 0.72, 1.22 |
| **Transportation mode** |  |  |  |  |  |  |  |  |
| Car-only | Ref. | - | Ref. | - | Ref. | - | Ref. | - |
| Public transport only | 1.14 | 1.09, 1.19 | 1.12 | 1.02, 1.23 | 1.05 | 0.99, 1.12 | 1.16 | 0.98, 1.36 |
| Active transport only | 0.87 | 0.83, 0.91 | 0.81 | 0.73, 0.89 | 0.89 | 0.84, 0.95 | 0.46 | 0.36, 0.58 |
| Mixed-without-car | 0.97 | 0.92, 1.01 | 0.96 | 0.87, 1.06 | 0.95 | 0.89, 1.01 | 0.81 | 0.67, 0.98 |
| Mixed-with-car | 0.88 | 0.86, 0.90 | 0.83 | 0.79, 0.88 | 0.93 | 0.90, 0.96 | 0.66 | 0.59, 0.74 |

Models adjusted for age, sex, ethnicity, education, Townsend deprivation index, annual household income, month of assessment, and population density.
Abbreviations: N = Number of participants; HR = Hazard ratio; CI = Confidence interval; CVD = cardiovascular disease; Ref. = Reference category

**Appendix Table 12.** Association of loneliness with all-cause and cause-specific mortality by non-commute transport subgroup, using car-only as the reference category and not adjusted for commuting transportation mode.

| Mortality outcome<br>N(deaths/total participants) | All-cause<br>40,410/468,292 |  | CVD<br>8,361/468,292 |  | Cancer<br>19,704/468,292 |  | Chronic respiratory<br>2,081/468,292 |  |
| --- | --- | --- | --- | --- | --- | --- | --- | --- |
|  | HR | 95% CI | HR | 95% CI | HR | 95% CI | HR | 95% CI |
| **Loneliness** |  |  |  |  |  |  |  |  |
| No | Ref. | - | Ref. | - | Ref. | - | Ref. | - |
| Yes | 1.19 | 1.15, 1.24 | 1.24 | 1.13, 1.35 | 1.05 | 0.99, 1.11 | 1.48 | 1.26, 1.73 |
| **Loneliness x transportation** |  |  |  |  |  |  |  |  |
| Yes x Public transport only | 0.99 | 0.92, 1.07 | 1.06 | 0.90, 1.24 | 0.96 | 0.85, 1.09 | 0.98 | 0.74, 1.29 |
| Yes x Active transport only | 0.99 | 0.90, 1.09 | 1.23 | 1.00, 1.50 | 1.00 | 0.87, 1.15 | 0.73 | 0.45, 1.19 |
| Yes x Mixed-without-car | 1.04 | 0.95, 1.13 | 1.17 | 0.98, 1.40 | 0.92 | 0.81, 1.06 | 1.01 | 0.71, 1.42 |
| Yes x Mixed-with-car | 0.97 | 0.91, 1.03 | 1.02 | 0.89, 1.16 | 1.01 | 0.93, 1.10 | 0.97 | 0.76, 1.25 |
| **Transportation mode** |  |  |  |  |  |  |  |  |
| Car-only | Ref. | - | Ref. | - | Ref. | - | Ref. | - |
| Public transport only | 1.16 | 1.11, 1.21 | 1.14 | 1.04, 1.25 | 1.07 | 1.01, 1.14 | 1.17 | 0.99, 1.39 |
| Active transport only | 0.86 | 0.83, 0.90 | 0.74 | 0.67, 0.82 | 0.89 | 0.83, 0.94 | 0.53 | 0.43, 0.67 |
| Mixed-without-car | 0.96 | 0.92, 1.00 | 0.93 | 0.85, 1.03 | 0.97 | 0.91, 1.04 | 0.77 | 0.64, 0.94 |
| Mixed-with-car | 0.87 | 0.85, 0.90 | 0.82 | 0.77, 0.87 | 0.92 | 0.89, 0.95 | 0.64 | 0.57, 0.73 |

Models adjusted for age, sex, ethnicity, education, Townsend deprivation index, annual household income, month of assessment, living alone status, and population density.
Abbreviations: N = Number of participants; HR = Hazard ratio; CI = Confidence interval; CVD = cardiovascular disease; Ref. = Reference category

**Appendix Table 13.** Association of social isolation with cause-specific mortality by non-commute transport subgroup, using Fine-Gray competing risks models with car-only as the reference category.

| Mortality outcome | CVD mortality |  | Cancer mortality |  | Respiratory mortality |  |
| --- | --- | --- | --- | --- | --- | --- |
| N(deaths/total population) | 8,232/455,072 |  | 19,317/455,072 |  | 2,072/455,072 |  |
|  | HR | 95% CI | HR | 95% CI | HR | 95% CI |
| **Social isolation** |  |  |  |  |  |  |
| No | Ref. | - | Ref. | - | Ref. | - |
| Yes | 1.41 | 1.27, 1.56 | 1.21 | 1.13, 1.30 | 1.59 | 1.32, 1.91 |
| **Social isolation x transportation mode** |  |  |  |  |  |  |
| Yes x Public transport only | 1.15 | 0.96, 1.38 | 0.99 | 0.86, 1.15 | 0.99 | 0.74, 1.33 |
| Yes x Active transport only | 0.89 | 0.73, 1.08 | 0.96 | 0.84, 1.10 | 1.19 | 0.76, 1.86 |
| Yes x Mixed-without-car | 1.11 | 0.95, 1.30 | 1.02 | 0.90, 1.15 | 0.87 | 0.61, 1.24 |
| Yes x Mixed-with-car | 0.95 | 0.82, 1.11 | 0.97 | 0.88, 1.06 | 0.99 | 0.73, 1.34 |
| **Transportation mode** |  |  |  |  |  |  |
| Car-only | Ref. | - | Ref. | - | Ref. | - |
| Public transport only | 1.11 | 1.01, 1.23 | 1.03 | 0.96, 1.11 | 1.11 | 0.94, 1.31 |
| Active transport only | 0.84 | 0.75, 0.93 | 0.90 | 0.85, 0.96 | 0.47 | 0.37, 0.60 |
| Mixed-without-car | 0.98 | 0.89, 1.08 | 0.94 | 0.90, 1.00 | 0.78 | 0.64, 0.96 |
| Mixed-with-car | 0.83 | 0.79, 0.88 | 0.93 | 0.89, 0.97 | 0.64 | 0.58, 0.71 |

Models adjusted for age, sex, ethnicity, education, Townsend deprivation index, annual household income, month of assessment, commute transportation mode, and population density.
Abbreviations: N = Number of participants; HR = Hazard ratio; CI = Confidence interval; CVD = cardiovascular disease; Ref. = Reference category

**Appendix Table 14.** Association of loneliness with cause-specific mortality by non-commute transport subgroup, using Fine-Gray competing risks models with car-only as the reference category.

| Mortality outcome | CVD mortality |  | Cancer mortality |  | Respiratory mortality |  |
| --- | --- | --- | --- | --- | --- | --- |
| N(deaths/total population) | 8,108/447,862 |  | 19,014/447,862 |  | 2,033/447,862 |  |
|  | HR | 95% CI | HR | 95% CI | HR | 95% CI |
| **Loneliness** |  |  |  |  |  |  |
| No | Ref. | - | Ref. | - | Ref. | - |
| Yes | 1.21 | 1.11, 1.31 | 1.04 | 0.98, 1.10 | 1.44 | 1.22, 1.70 |
| **Loneliness x transportation** |  |  |  |  |  |  |
| Yes x Public transport only | 1.06 | 0.90, 1.26 | 0.95 | 0.83, 1.08 | 0.96 | 0.73, 1.27 |
| Yes x Active transport only | **1.24** | **1.04, 1.48** | 0.99 | 0.84, 1.16 | 0.72 | 0.45, 1.15 |
| Yes x Mixed-without-car | 1.16 | 0.97, 1.38 | 0.90 | 0.79, 1.04 | 0.99 | 0.69, 1.40 |
| Yes x Mixed-with-car | 1.04 | 0.91, 1.18 | 1.01 | 0.93, 1.10 | 1.00 | 0.79, 1.26 |
| **Transportation mode** |  |  |  |  |  |  |
| Car-only | Ref. | - | Ref. | - | Ref. | - |
| Public transport only | 1.12 | 1.03, 1.22 | 1.05 | 0.99, 1.12 | 1.11 | 0.92, 1.34 |
| Active transport only | 0.76 | 0.70, 0.83 | 0.90 | 0.84, 0.96 | 0.55 | 0.44, 0.69 |
| Mixed-without-car | 0.95 | 0.86, 1.06 | 0.97 | 0.91, 1.03 | 0.75 | 0.60, 0.93 |
| Mixed-with-car | 0.82 | 0.77, 0.87 | 0.92 | 0.89, 0.96 | 0.63 | 0.55, 0.73 |

Models adjusted for age, sex, ethnicity, education, Townsend deprivation index, annual household income, month of assessment, commute transportation mode, living alone status, and population density.
Abbreviations: N = Number of participants; HR = Hazard ratio; CI = Confidence interval; CVD = cardiovascular disease; Ref. = Reference category

**Appendix Table 15.**
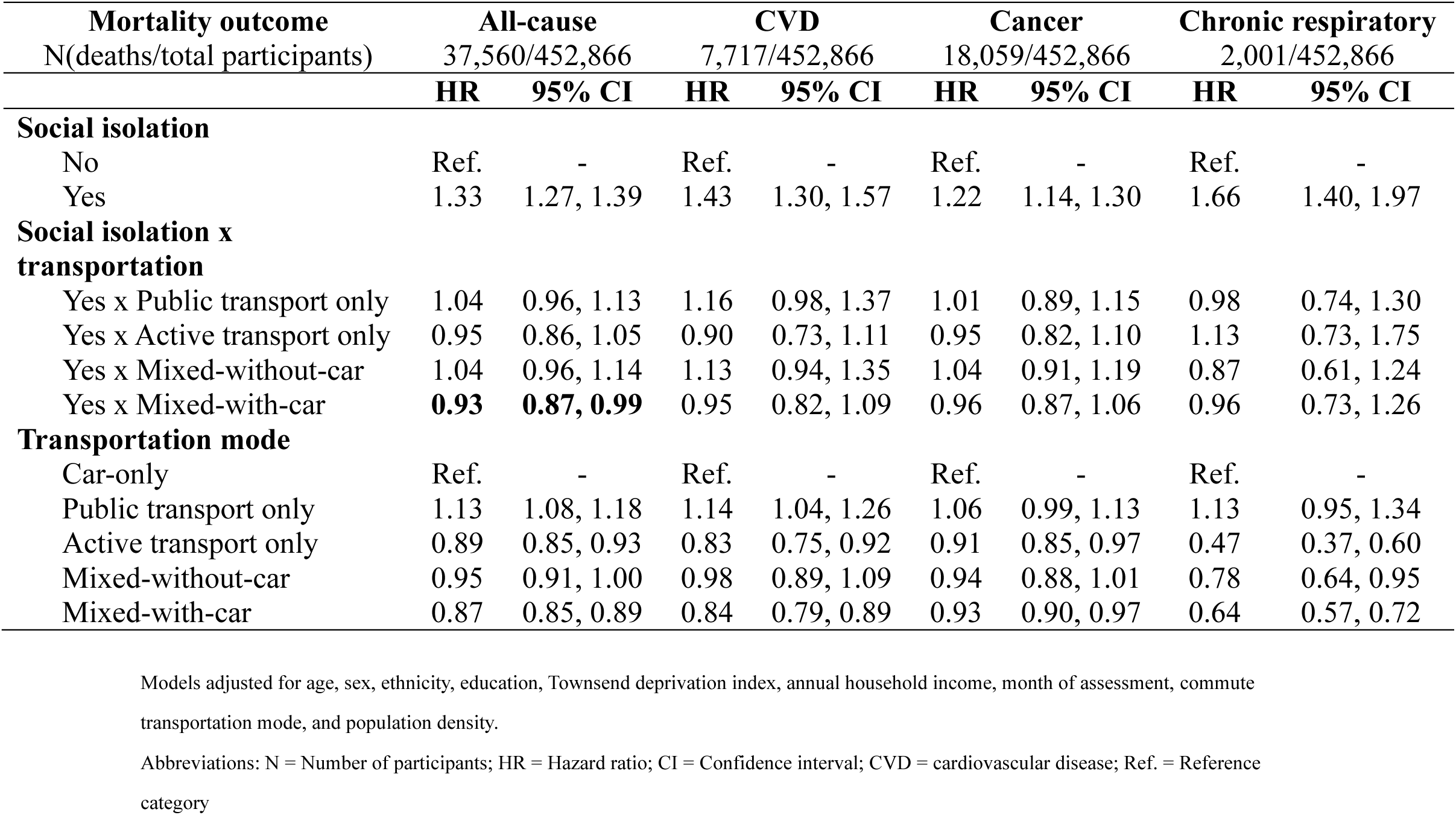
Association of social isolation with all-cause and cause-specific mortality by non-commute transport subgroup, using car-only as the reference category and excluding events that occurred within two years of baseline.

**Appendix Table 16.** Association of loneliness with all-cause and cause-specific mortality by non-commute transport subgroup, using car-only as the reference category and excluding events that occurred within two years of baseline.

| Mortality outcome<br>N(deaths/total participants) | All-cause<br>36,943/445,692 |  | CVD<br>7,599/445,692 |  | Cancer<br>17,779/445,692 |  | Chronic respiratory<br>1,962/445,692 |  |
| --- | --- | --- | --- | --- | --- | --- | --- | --- |
|  | HR | 95% CI | HR | 95% CI | HR | 95% CI | HR | 95% CI |
| **Loneliness** |  |  |  |  |  |  |  |  |
| No | Ref. | - | Ref. | - | Ref. | - | Ref. | - |
| Yes | 1.19 | 1.14, 1.24 | 1.22 | 1.12, 1.34 | 1.04 | 0.97, 1.10 | 1.47 | 1.25, 1.73 |
| **Loneliness x transportation** |  |  |  |  |  |  |  |  |
| Yes x Public transport only | 0.99 | 0.91, 1.07 | 1.09 | 0.92, 1.29 | 0.99 | 0.87, 1.12 | 0.96 | 0.73, 1.28 |
| Yes x Active transport only | 0.98 | 0.89, 1.09 | **1.25** | **1.02, 1.55** | 1.00 | 0.86, 1.17 | 0.67 | 0.40, 1.10 |
| Yes x Mixed-without-car | 1.05 | 0.96, 1.14 | 1.16 | 0.96, 1.39 | 0.94 | 0.81, 1.08 | 0.99 | 0.69, 1.41 |
| Yes x Mixed-with-car | 0.98 | 0.92, 1.04 | 1.02 | 0.89, 1.17 | 1.04 | 0.95, 1.13 | 0.97 | 0.75, 1.25 |
| **Transportation mode** |  |  |  |  |  |  |  |  |
| Car-only | Ref. | - | Ref. | - | Ref. | - | Ref. | - |
| Public transport only | 1.14 | 1.09, 1.20 | 1.14 | 1.04, 1.26 | 1.07 | 1.00, 1.14 | 1.13 | 0.95, 1.34 |
| Active transport only | 0.88 | 0.84, 0.92 | 0.76 | 0.68, 0.85 | 0.90 | 0.84, 0.96 | 0.55 | 0.44, 0.69 |
| Mixed-without-car | 0.95 | 0.90, 0.99 | 0.96 | 0.87, 1.06 | 0.96 | 0.90, 1.03 | 0.74 | 0.60, 0.91 |
| Mixed-with-car | 0.86 | 0.84, 0.89 | 0.83 | 0.78, 0.88 | 0.92 | 0.89, 0.96 | 0.63 | 0.56, 0.72 |

Models adjusted for age, sex, ethnicity, education, Townsend deprivation index, annual household income, month of assessment, commute transportation mode, living alone status, and population density.
Abbreviations: N = Number of participants; HR = Hazard ratio; CI = Confidence interval; CVD = cardiovascular disease; Ref. = Reference category

**Appendix Table 17.**
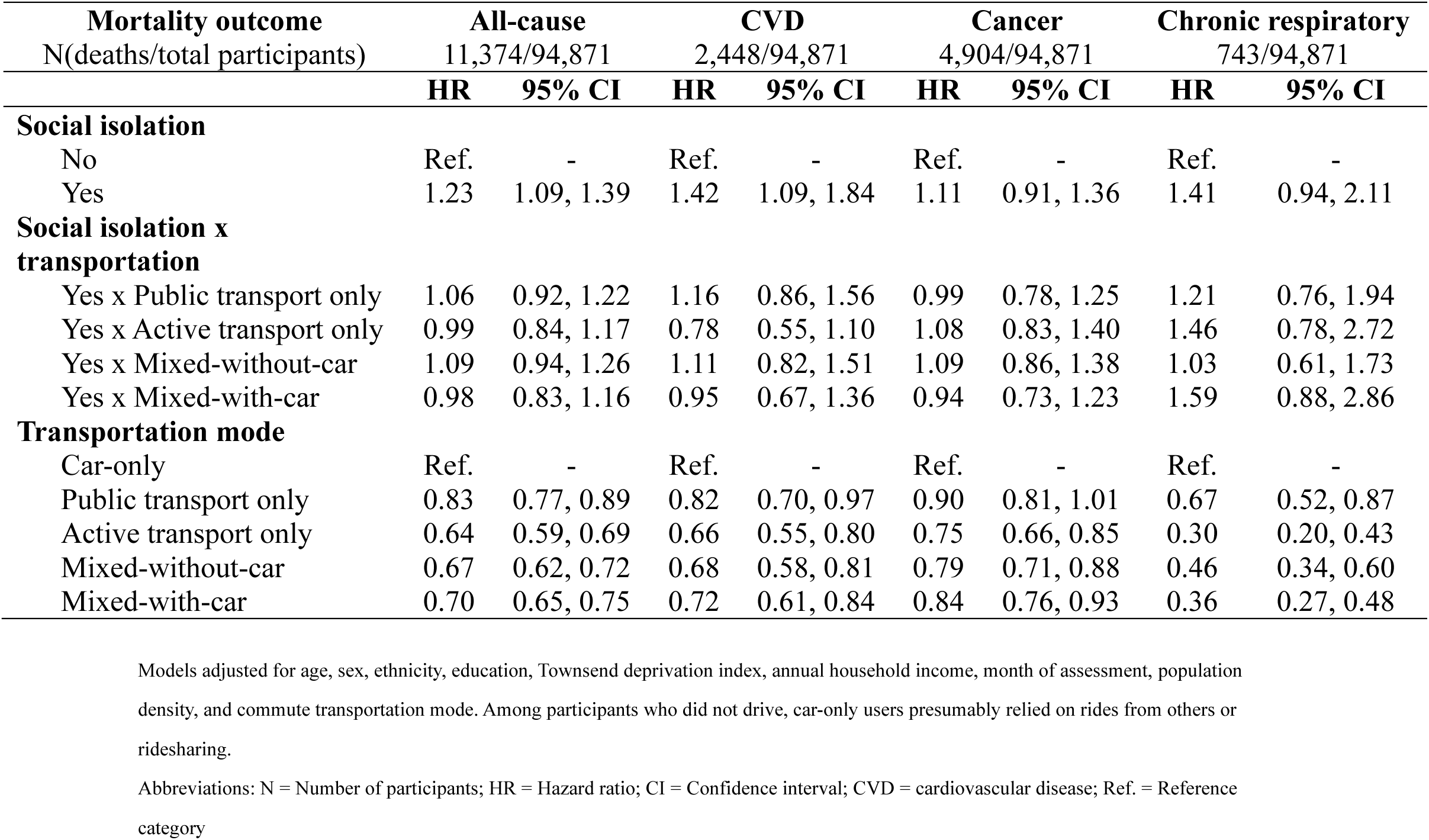
Association of social isolation with all-cause and cause-specific mortality by non-commute transport subgroup, using car-only as the reference category and restricting to participants who did not drive.

**Appendix Table 18.** Association of loneliness with all-cause and cause-specific mortality by non-commute transport subgroup, using car-only as the reference category and restricting to participants who did not drive.

| Mortality outcome<br>N(deaths/total participants) | All-cause<br>11,135/92,913 |  | CVD<br>2,395/92,913 |  | Cancer<br>4,812/92,913 |  | Chronic respiratory<br>731/92,913 |  |
| --- | --- | --- | --- | --- | --- | --- | --- | --- |
|  | HR | 95% CI | HR | 95% CI | HR | 95% CI | HR | 95% CI |
| **Loneliness** |  |  |  |  |  |  |  |  |
| No | Ref. | - | Ref. | - | Ref. | - | Ref. | - |
| Yes | 1.30 | 1.16, 1.45 | 1.21 | 0.94, 1.56 | 1.07 | 0.89, 1.29 | 2.06 | 1.46, 2.92 |
| **Loneliness x transportation** |  |  |  |  |  |  |  |  |
| Yes x Public transport only | **0.83** | **0.73, 0.95** | 0.95 | 0.71, 1.28 | 0.90 | 0.72, 1.12 | **0.66** | **0.43, 1.00** |
| Yes x Active transport only | 0.87 | 0.74, 1.02 | 1.36 | 0.97, 1.92 | 0.94 | 0.72, 1.21 | **0.52** | **0.27, 0.98** |
| Yes x Mixed-without-car | 0.95 | 0.82, 1.09 | 1.13 | 0.83, 1.53 | 0.93 | 0.74, 1.17 | 0.66 | 0.41, 1.08 |
| Yes x Mixed-with-car | 0.92 | 0.80, 1.07 | 1.18 | 0.85, 1.64 | 0.99 | 0.78, 1.25 | 0.82 | 0.48, 1.41 |
| **Transportation mode** |  |  |  |  |  |  |  |  |
| Car-only | Ref. | - | Ref. | - | Ref. | - | Ref. | - |
| Public transport only | 0.85 | 0.79, 0.92 | 0.83 | 0.71, 0.98 | 0.90 | 0.81, 1.02 | 0.76 | 0.58, 1.01 |
| Active transport only | 0.64 | 0.59, 0.70 | 0.53 | 0.44, 0.64 | 0.76 | 0.67, 0.87 | 0.41 | 0.28, 0.58 |
| Mixed-without-car | 0.67 | 0.62, 0.72 | 0.64 | 0.54, 0.76 | 0.80 | 0.71, 0.90 | 0.48 | 0.36, 0.65 |
| Mixed-with-car | 0.71 | 0.66, 0.76 | 0.67 | 0.57, 0.79 | 0.84 | 0.75, 0.94 | 0.42 | 0.31, 0.57 |

Models adjusted for age, sex, ethnicity, education, Townsend deprivation index, annual household income, month of assessment, commute transportation mode, living alone status, population density, and commute transportation mode. Among participants who did not drive, car-only users presumably relied on rides from others or ridesharing.
Abbreviations: N = Number of participants; HR = Hazard ratio; CI = Confidence interval; CVD = cardiovascular disease; Ref. = Reference category

**Appendix Table 19.** Association of social isolation with all-cause and cause-specific mortality by non-commute transport subgroup, using car-only as the reference category and restricting to urban participants only.

| Mortality outcome<br>N(deaths/total participants) | All-cause<br>34,307/387,383 |  | CVD<br>7,083/387,383 |  | Cancer<br>16,580/387,383 |  | Chronic respiratory<br>1,829/387,383 |  |
| --- | --- | --- | --- | --- | --- | --- | --- | --- |
|  | HR | 95% CI | HR | 95% CI | HR | 95% CI | HR | 95% CI |
| **Social isolation** |  |  |  |  |  |  |  |  |
| No | Ref. | - | Ref. | - | Ref. | - | Ref. | - |
| Yes | 1.32 | 1.26, 1.39 | 1.45 | 1.31, 1.60 | 1.22 | 1.14, 1.31 | 1.49 | 1.24, 1.80 |
| **Social isolation x transportation** |  |  |  |  |  |  |  |  |
| Yes x Public transport only | 1.07 | 0.98, 1.16 | 1.14 | 0.97, 1.35 | 1.02 | 0.90, 1.16 | 1.10 | 0.82, 1.48 |
| Yes x Active transport only | 0.97 | 0.88, 1.07 | 0.89 | 0.72, 1.10 | 0.97 | 0.84, 1.12 | 1.27 | 0.82, 1.98 |
| Yes x Mixed-without-car | 1.04 | 0.95, 1.13 | 1.10 | 0.91, 1.32 | 1.03 | 0.90, 1.18 | 0.93 | 0.64, 1.34 |
| Yes x Mixed-with-car | 0.94 | 0.88, 1.01 | 0.91 | 0.78, 1.06 | 0.97 | 0.87, 1.07 | 1.09 | 0.82, 1.46 |
| **Transportation mode** |  |  |  |  |  |  |  |  |
| Car-only | Ref. | - | Ref. | - | Ref. | - | Ref. | - |
| Public transport only | 1.12 | 1.07, 1.18 | 1.12 | 1.02, 1.24 | 1.02 | 0.96, 1.10 | 1.13 | 0.95, 1.35 |
| Active transport only | 0.87 | 0.83, 0.91 | 0.8 | 0.72, 0.90 | 0.89 | 0.83, 0.95 | 0.49 | 0.38, 0.63 |
| Mixed-without-car | 0.94 | 0.90, 0.99 | 0.97 | 0.87, 1.07 | 0.92 | 0.86, 0.99 | 0.78 | 0.63, 0.95 |
| Mixed-with-car | 0.86 | 0.84, 0.88 | 0.81 | 0.76, 0.86 | 0.92 | 0.89, 0.96 | 0.62 | 0.55, 0.71 |

Models adjusted for age, sex, ethnicity, education, Townsend deprivation index, annual household income, month of assessment, and commute transportation mode.
Abbreviations: N = Number of participants; HR = Hazard ratio; CI = Confidence interval; CVD = cardiovascular disease; Ref. = Reference category

**Appendix Table 20.** Association of loneliness with all-cause and cause-specific mortality by non-commute transport subgroup, using car-only as the reference category and restricting to urban participants only.

| Mortality outcome<br>N(deaths/total participants) | All-cause<br>33,720/381,031 |  | CVD<br>6,974/381,031 |  | Cancer<br>16,307/381,031 |  | Chronic respiratory<br>1,795/381,031 |  |
| --- | --- | --- | --- | --- | --- | --- | --- | --- |
|  | HR | 95% CI | HR | 95% CI | HR | 95% CI | HR | 95% CI |
| **Loneliness** |  |  |  |  |  |  |  |  |
| No | Ref. | - | Ref. | - | Ref. | - | Ref. | - |
| Yes | 1.19 | 1.14, 1.24 | 1.22 | 1.11, 1.34 | 1.04 | 0.97, 1.11 | 1.50 | 1.26, 1.78 |
| **Loneliness x transportation** |  |  |  |  |  |  |  |  |
| Yes x Public transport only | 0.96 | 0.89, 1.04 | 1.05 | 0.89, 1.24 | 0.93 | 0.81, 1.06 | 0.91 | 0.69, 1.21 |
| Yes x Active transport only | 0.96 | 0.87, 1.07 | 1.17 | 0.94, 1.46 | 0.99 | 0.85, 1.15 | 0.68 | 0.41, 1.12 |
| Yes x Mixed-without-car | 1.04 | 0.95, 1.13 | 1.15 | 0.95, 1.38 | 0.93 | 0.81, 1.07 | 1.00 | 0.70, 1.43 |
| Yes x Mixed-with-car | 0.97 | 0.91, 1.04 | 1.03 | 0.90, 1.19 | 1.01 | 0.92, 1.11 | 1.01 | 0.77, 1.32 |
| **Transportation mode** |  |  |  |  |  |  |  |  |
| Car-only | Ref. | - | Ref. | - | Ref. | - | Ref. | - |
| Public transport only | 1.15 | 1.10, 1.20 | 1.14 | 1.03, 1.26 | 1.06 | 0.99, 1.13 | 1.19 | 1.00, 1.42 |
| Active transport only | 0.87 | 0.83, 0.91 | 0.75 | 0.67, 0.83 | 0.88 | 0.82, 0.94 | 0.58 | 0.46, 0.74 |
| Mixed-without-car | 0.93 | 0.89, 0.98 | 0.94 | 0.85, 1.04 | 0.94 | 0.88, 1.01 | 0.74 | 0.60, 0.91 |
| Mixed-with-car | 0.86 | 0.83, 0.88 | 0.79 | 0.74, 0.84 | 0.92 | 0.88, 0.96 | 0.62 | 0.55, 0.71 |

Models adjusted for age, sex, ethnicity, education, Townsend deprivation index, annual household income, month of assessment, commute transportation mode, and living alone status.
Abbreviations: N = Number of participants; HR = Hazard ratio; CI = Confidence interval; CVD = cardiovascular disease; Ref. = Reference category

**Appendix Table 21.** Association of social isolation with all-cause and cause-specific mortality by non-commute transport subgroup, using car-only as the reference category and excluding participants who selected multiple modes.

| Mortality outcome<br>N(deaths/total participants) | All-cause<br>22,126/244,659 |  | CVD<br>4,667/244,659 |  | Cancer<br>10,402/244,659 |  | Chronic respiratory<br>1,280/244,659 |  |
| --- | --- | --- | --- | --- | --- | --- | --- | --- |
|  | HR | 95% CI | HR | 95% CI | HR | 95% CI | HR | 95% CI |
| **Social isolation** |  |  |  |  |  |  |  |  |
| No | Ref. | - | Ref. | - | Ref. | - | Ref. | - |
| Yes | 1.33 | 1.28, 1.39 | 1.44 | 1.31, 1.57 | 1.23 | 1.15, 1.31 | 1.66 | 1.40, 1.96 |
| **Social isolation x transportation** |  |  |  |  |  |  |  |  |
| Yes x Public transport only | 1.05 | 0.97, 1.13 | 1.17 | 0.99, 1.37 | 1.01 | 0.89, 1.14 | 0.99 | 0.75, 1.31 |
| Yes x Active transport only | 0.94 | 0.85, 1.03 | 0.88 | 0.71, 1.08 | 0.95 | 0.83, 1.09 | 1.16 | 0.76, 1.78 |
| **Transportation mode** |  |  |  |  |  |  |  |  |
| Car-only | Ref. | - | Ref. | - | Ref. | - | Ref. | - |
| Public transport only | 1.11 | 1.06, 1.16 | 1.11 | 1.00, 1.22 | 1.02 | 0.96, 1.09 | 1.10 | 0.92, 1.30 |
| Active transport only | 0.87 | 0.83, 0.91 | 0.83 | 0.75, 0.91 | 0.89 | 0.84, 0.95 | 0.46 | 0.36, 0.59 |

Models adjusted for age, sex, ethnicity, education, Townsend deprivation index, annual household income, month of assessment, commute transportation mode, and population density. Abbreviations: N = Number of participants; HR = Hazard ratio; CI = Confidence interval; CVD = cardiovascular disease; Ref. = Reference category

**Appendix Table 22.** Association of loneliness with all-cause and cause-specific mortality by non-commute transport subgroup, using car-only as the reference category and excluding participants who selected multiple modes.

| Mortality outcome<br>N(deaths/total participants) | All-cause<br>21,759/240,781 |  | CVD<br>4,600/240,781 |  | Cancer<br>10,234/240,781 |  | Chronic respiratory<br>1,262/240,781 |  |
| --- | --- | --- | --- | --- | --- | --- | --- | --- |
|  | HR | 95% CI | HR | 95% CI | HR | 95% CI | HR | 95% CI |
| **Loneliness** |  |  |  |  |  |  |  |  |
| No | Ref. | - | Ref. | - | Ref. | - | Ref. | - |
| Yes | 1.18 | 1.13, 1.23 | 1.21 | 1.11, 1.33 | 1.05 | 0.99, 1.12 | 1.42 | 1.21, 1.68 |
| **Loneliness x transportation** |  |  |  |  |  |  |  |  |
| Yes x Public transport only | 0.97 | 0.90, 1.05 | 1.05 | 0.90, 1.24 | 0.95 | 0.84, 1.07 | 0.95 | 0.72, 1.25 |
| Yes x Active transport only | 0.98 | 0.89, 1.08 | **1.23** | **1.00, 1.51** | 0.98 | 0.85, 1.14 | 0.72 | 0.44, 1.16 |
| **Transportation mode** |  |  |  |  |  |  |  |  |
| Car-only | Ref. | - | Ref. | - | Ref. | - | Ref. | - |
| Public transport only | 1.12 | 1.07, 1.17 | 1.12 | 1.02, 1.24 | 1.04 | 0.97, 1.12 | 1.10 | 0.93, 1.32 |
| Active transport only | 0.86 | 0.82, 0.90 | 0.75 | 0.68, 0.83 | 0.89 | 0.83, 0.95 | 0.54 | 0.43, 0.67 |

Models adjusted for age, sex, ethnicity, education, Townsend deprivation index, annual household income, month of assessment, commute transportation mode, living alone status, and population density. Abbreviations: N = Number of participants; HR = Hazard ratio; CI = Confidence interval; CVD = cardiovascular disease; Ref. = Reference category

**Appendix Table 23.** Association of social isolation with all-cause and cause-specific mortality by non-commute transport subgroup, using car-only as the reference category and restricting to female participants only.

| Mortality outcome<br>N(deaths/total participants) | All-cause<br>16,515/250,085 |  | CVD<br>2,572 /250,085 |  | Cancer<br>9,068/250,085 |  | Chronic respiratory<br>789/250,085 |  |
| --- | --- | --- | --- | --- | --- | --- | --- | --- |
|  | HR | 95% CI | HR | 95% CI | HR | 95% CI | HR | 95% CI |
| **Social isolation** |  |  |  |  |  |  |  |  |
| No | Ref. | - | Ref. | - | Ref. | - | Ref. | - |
| Yes | 1.34 | 1.25, 1.44 | 1.46 | 1.23, 1.74 | 1.26 | 1.14 1.39 | 1.64 | 1.24, 2.17 |
| **Social isolation x transportation** |  |  |  |  |  |  |  |  |
| Yes x Public transport only | 1.04 | 0.92, 1.17 | 1.08 | 0.81, 1.45 | 0.98 | 0.82, 1.18 | 1.31 | 0.85, 2.03 |
| Yes x Active transport only | 0.99 | 0.84, 1.16 | 0.87 | 0.57, 1.33 | 1.02 | 0.82, 1.28 | 1.02 | 0.49, 2.15 |
| Yes x Mixed-without-car | 1.03 | 0.90, 1.18 | 1.04 | 0.76, 1.42 | 1.05 | 0.87, 1.27 | 0.72 | 0.41, 1.26 |
| Yes x Mixed-with-car | 0.94 | 0.85, 1.05 | 1.01 | 0.79, 1.30 | 0.95 | 0.82, 1.09 | 1.30 | 0.83, 2.02 |
| **Transportation mode** |  |  |  |  |  |  |  |  |
| Car-only | Ref. | - | Ref. | - | Ref. | - | Ref. | - |
| Public transport only | 1.02 | 0.96, 1.09 | 1.00 | 0.85, 1.17 | 0.99 | 0.91, 1.09 | 0.89 | 0.68, 1.15 |
| Active transport only | 0.79 | 0.74, 0.85 | 0.78 | 0.64, 0.93 | 0.81 | 0.74, 0.90 | 0.50 | 0.35, 0.73 |
| Mixed-without-car | 0.88 | 0.82, 0.94 | 0.89 | 0.75, 1.05 | 0.90 | 0.82, 0.99 | 0.78 | 0.59, 1.04 |
| Mixed-with-car | 0.85 | 0.82, 0.89 | 0.78 | 0.70, 0.86 | 0.94 | 0.89, 0.99 | 0.48 | 0.40, 0.59 |

Models adjusted for age, ethnicity, education, Townsend deprivation index, annual household income, month of assessment, commute transportation mode, and population density.
Abbreviations: N = Number of participants; HR = Hazard ratio; CI = Confidence interval; CVD = cardiovascular disease; Ref. = Reference category

**Appendix Table 24.** Association of social isolation with all-cause and cause-specific mortality by non-commute transport subgroup, using car-only as the reference category and restricting to male participants only.

| Mortality outcome<br>N(deaths/total participants) | All-cause<br>23,251/204,987 |  | CVD<br>5,660/204,987 |  | Cancer<br>10,249/204,987 |  | Chronic respiratory<br>1,283/204,987 |  |
| --- | --- | --- | --- | --- | --- | --- | --- | --- |
|  | HR | 95% CI | HR | 95% CI | HR | 95% CI | HR | 95% CI |
| **Social isolation** |  |  |  |  |  |  |  |  |
| No | Ref. | - | Ref. | - | Ref. | - | Ref. | - |
| Yes | 1.35 | 1.28, 1.42 | 1.43 | 1.29, 1.60 | 1.22 | 1.12, 1.33 | 1.66 | 1.34, 2.05 |
| **Social isolation x transportation** |  |  |  |  |  |  |  |  |
| Yes x Public transport only | 1.04 | 0.94, 1.15 | 1.19 | 0.98, 1.45 | 1.01 | 0.86, 1.20 | 0.82 | 0.57, 1.18 |
| Yes x Active transport only | 0.91 | 0.81, 1.02 | 0.87 | 0.69, 1.11 | 0.91 | 0.76, 1.08 | 1.29 | 0.76, 2.18 |
| Yes x Mixed-without-car | 1.02 | 0.91, 1.13 | 1.14 | 0.92, 1.40 | 0.98 | 0.82, 1.17 | 1.01 | 0.65, 1.59 |
| Yes x Mixed-with-car | **0.92** | **0.85, 1.00** | 0.91 | 0.77, 1.08 | 0.97 | 0.86, 1.10 | 0.83 | 0.59, 1.17 |
| **Transportation mode** |  |  |  |  |  |  |  |  |
| Car-only | Ref. | - | Ref. | - | Ref. | - | Ref. | - |
| Public transport only | 1.24 | 1.17, 1.31 | 1.19 | 1.06, 1.34 | 1.12 | 1.02, 1.23 | 1.32 | 1.06, 1.64 |
| Active transport only | 0.94 | 0.88, 0.99 | 0.85 | 0.76, 0.96 | 0.96 | 0.89, 1.05 | 0.44 | 0.32, 0.61 |
| Mixed-without-car | 1.02 | 0.96, 1.09 | 1.02 | 0.90, 1.15 | 0.99 | 0.90, 1.09 | 0.75 | 0.57, 0.98 |
| Mixed-with-car | 0.87 | 0.84, 0.90 | 0.85 | 0.79, 0.90 | 0.91 | 0.87, 0.96 | 0.73 | 0.63, 0.85 |

Models adjusted for age, ethnicity, education, Townsend deprivation index, annual household income, month of assessment, commute transportation mode, and population density.
Abbreviations: N = Number of participants; HR = Hazard ratio; CI = Confidence interval; CVD = cardiovascular disease; Ref. = Reference category

**Appendix Table 25.** Association of loneliness with all-cause and cause-specific mortality by non-commute transport subgroup, using car-only as the reference category and restricting to female participants only.

| Mortality outcome<br>N(deaths/total participants) | All-cause<br>16,227/245,945 |  | CVD<br>2,536/245,945 |  | Cancer<br>8,913/245,945 |  | Chronic respiratory<br>770/245,945 |  |
| --- | --- | --- | --- | --- | --- | --- | --- | --- |
|  | HR | 95% CI | HR | 95% CI | HR | 95% CI | HR | 95% CI |
| **Loneliness** |  |  |  |  |  |  |  |  |
| No | Ref. | - | Ref. | - | Ref. | - | Ref. | - |
| Yes | 1.18 | 1.11, 1.25 | 1.23 | 1.06, 1.43 | 1.03 | 0.95, 1.12 | 1.69 | 1.33, 2.14 |
| **Loneliness x transportation** |  |  |  |  |  |  |  |  |
| Yes x Public transport only | 1.01 | 0.90, 1.13 | 1.10 | 0.84, 1.44 | 1.03 | 0.87, 1.22 | 0.85 | 0.56, 1.28 |
| Yes x Active transport only | 0.98 | 0.84, 1.14 | 1.25 | 0.87, 1.81 | 1.00 | 0.81, 1.24 | 0.54 | 0.25, 1.17 |
| Yes x Mixed-without-car | 0.98 | 0.86, 1.12 | 1.21 | 0.90, 1.63 | 0.90 | 0.75, 1.09 | 0.89 | 0.54, 1.47 |
| Yes x Mixed-with-car | 0.97 | 0.89, 1.06 | 0.99 | 0.80, 1.23 | 1.04 | 0.93, 1.17 | 0.88 | 0.60, 1.31 |
| **Transportation mode** |  |  |  |  |  |  |  |  |
| Car-only | Ref. | - | Ref. | - | Ref. | - | Ref. | - |
| Public transport only | 1.03 | 0.96, 1.10 | 1.00 | 0.84, 1.18 | 0.98 | 0.89, 1.08 | 1.02 | 0.78, 1.35 |
| Active transport only | 0.79 | 0.74, 0.86 | 0.71 | 0.58, 0.87 | 0.81 | 0.74, 0.90 | 0.60 | 0.41, 0.87 |
| Mixed-without-car | 0.89 | 0.83, 0.95 | 0.85 | 0.71, 1.01 | 0.94 | 0.85, 1.03 | 0.73 | 0.54, 1.00 |
| Mixed-with-car | 0.85 | 0.82, 0.89 | 0.78 | 0.70, 0.87 | 0.92 | 0.88, 0.98 | 0.51 | 0.41, 0.63 |

Models adjusted for age, ethnicity, education, Townsend deprivation index, annual household income, month of assessment, commute transportation mode, living alone status, and population density.
Abbreviations: N = Number of participants; HR = Hazard ratio; CI = Confidence interval; CVD = cardiovascular disease; Ref. = Reference category

**Appendix Table 26.** Association of loneliness with all-cause and cause-specific mortality by non-commute transport subgroup, using car-only as the reference category and restricting to male participants only.

| Mortality outcome<br>N(deaths/total participants) | All-cause<br>22,827/201,696 |  | CVD<br>5,552/201,696 |  | Cancer<br>10,083/201,696 |  | Chronic respiratory<br>1,260/201,696 |  |
| --- | --- | --- | --- | --- | --- | --- | --- | --- |
|  | HR | 95% CI | HR | 95% CI | HR | 95% CI | HR | 95% CI |
| **Loneliness** |  |  |  |  |  |  |  |  |
| No | Ref. | - | Ref. | - | Ref. | - | Ref. | - |
| Yes | 1.19 | 1.13, 1.26 | 1.20 | 1.08, 1.35 | 1.07 | 0.98, 1.17 | 1.26 | 1.01, 1.58 |
| **Loneliness x transportation** |  |  |  |  |  |  |  |  |
| Yes x Public transport only | 0.97 | 0.87, 1.08 | 1.04 | 0.85, 1.27 | 0.9 | 0.75, 1.08 | 1.06 | 0.73, 1.54 |
| Yes x Active transport only | 1.00 | 0.88, 1.13 | 1.22 | 0.96, 1.56 | 1.00 | 0.82, 1.22 | 0.89 | 0.47, 1.66 |
| Yes x Mixed-without-car | 1.11 | 0.98, 1.24 | 1.15 | 0.92, 1.44 | 0.94 | 0.77, 1.15 | 1.10 | 0.68, 1.79 |
| Yes x Mixed-with-car | 0.98 | 0.90, 1.06 | 1.06 | 0.90, 1.25 | 0.97 | 0.85, 1.11 | 1.18 | 0.84, 1.65 |
| **Transportation mode** |  |  |  |  |  |  |  |  |
| Car-only | Ref. | - | Ref. | - | Ref. | - | Ref. | - |
| Public transport only | 1.25 | 1.18, 1.32 | 1.21 | 1.08, 1.36 | 1.16 | 1.06, 1.27 | 1.21 | 0.97, 1.50 |
| Active transport only | 0.91 | 0.86, 0.96 | 0.77 | 0.68, 0.87 | 0.95 | 0.87, 1.03 | 0.51 | 0.39, 0.68 |
| Mixed-without-car | 0.99 | 0.93, 1.05 | 0.99 | 0.87, 1.11 | 1.00 | 0.91, 1.10 | 0.73 | 0.56, 0.94 |
| Mixed-with-car | 0.86 | 0.83, 0.89 | 0.82 | 0.76, 0.87 | 0.91 | 0.87, 0.96 | 0.69 | 0.59, 0.79 |

Models adjusted for age, ethnicity, education, Townsend deprivation index, annual household income, month of assessment, commute transportation mode, living alone status, and population density.
Abbreviations: N = Number of participants; HR = Hazard ratio; CI = Confidence interval; CVD = cardiovascular disease; Ref. = Reference category

**Appendix Table 27.** Characteristics of participants with complete baseline social isolation data compared to participants excluded due to missing social isolation data.

|  | Complete baseline social isolation<sup>1</sup><br>N = 490,114 | Excluded (missing baseline social isolation)<sup>1</sup><br>N = 10,709 |
| --- | --- | --- |
| Death | 42,913 (8.8%) | 1,581 (15%) |
| Loneliness (Yes) | 88,819 (18%) | 2,245 (25%) |
| Unknown | 8,154 | 1,667 |
| Age | 58.3 (50.6, 63.7) | 58.5 (50.4, 64.3) |
| Sex (Female) | 266,967 (54%) | 5,490 (51%) |
| Education |  |  |
| O levels/GCSEs or equivalent, CSEs or equivalent | 82,195 (17%) | 876 (17%) |
| A levels/AS levels, NVQ/HND/HNC or equivalent | 160,748 (33%) | 1,423 (27%) |
| College or University degree | 159,350 (33%) | 1,033 (20%) |
| None of the above | 83,220 (17%) | 1,900 (36%) |
| Unknown | 4,601 | 5,477 |
| Household income |  |  |
| Less than £18,000 | 96,035 (20%) | 918 (19%) |
| £18,000 to £30,999 | 107,315 (22%) | 562 (12%) |
| £31,000 to £51,999 | 110,014 (22%) | 409 (8.6%) |
| Greater than £52,000 | 108,444 (22%) | 253 (5.3%) |
| Do not know/ | 68,306 (14%) | 2,591 (55%) |
| Prefer not to answer |  |  |
| Unknown | 0 | 5,976 |
| Live alone | 90,893 (19%) | 1,694 (20%) |
| Unknown | 0 | 2,242 |
| Ethnicity |  |  |
| Non-white | 25,559 (5.2%) | 1,318 (14%) |
| White | 462,952 (95%) | 8,239 (86%) |
| Unknown | 1,603 | 1,152 |
| Home Population Density |  |  |
| Urban | 412,365 (85%) | 10,017 (95%) |
| Non-urban | 72,955 (15%) | 578 (5.5%) |
| Unknown | 4,794 | 114 |
| Townsend Deprivation Index | -2.2 (-3.7, 0.5) | -0.3 (-2.8, 3.3) |
| Unknown | 612 | 10 |
| Alcohol consumption |  |  |
| Frequent drinking | 213,312 (44%) | 3,163 (33%) |
| Occasional drinking | 181,299 (37%) | 3,270 (34%) |
| Special occasions only | 56,158 (11%) | 1,658 (17%) |
| Never | 38,957 (8.0%) | 1,521 (16%) |
| Unknown | 388 | 1,097 |
| Smoking |  |  |
| Current | 51,310 (11%) | 1,474 (15%) |
| Never | 267,575 (55%) | 5,093 (53%) |
| Previous | 169,475 (35%) | 2,969 (31%) |
| Unknown | 1,754 | 1,173 |

|  | Complete baseline social<br>isolation<sup>1</sup><br>N = 490,114 | Excluded (missing baseline<br>social isolation)<sup>1</sup><br>N = 10,709 |
| --- | --- | --- |
| Sleep duration (hours) | 7.0 (6.0, 8.0) | 7.0 (6.0, 8.0) |
| Unknown | 4 | 871 |
| MVPA self-reported | 315.0 (70.0, 630.0) | 0.0 (0.0, 98.0) |
| Unknown | 19,235 | 172 |
| Red meat |  |  |
| More than 4 times a week | 242,863 (50%) | 4,490 (47%) |
| 2-4 times a week | 154,877 (32%) | 2,666 (28%) |
| Less than 2 times a week | 91,676 (19%) | 2,417 (25%) |
| Unknown | 698 | 1,136 |
| Processed meat |  |  |
| More than 2 times a week | 151,896 (31%) | 2,780 (29%) |
| Less than 2 times a week | 337,263 (69%) | 6,673 (71%) |
| Unknown | 955 | 1,256 |
| Cooked vegetables |  |  |
| More than 4 servings/day | 49,528 (10%) | 1,132 (13%) |
| 2 to 4 servings/day | 346,340 (72%) | 5,796 (65%) |
| < 2 servings/day | 88,444 (18%) | 2,029 (23%) |
| Unknown | 5,802 | 1,752 |
| Oily fish |  |  |
| More than 2 times a week | 87,898 (18%) | 1,973 (21%) |
| Less than 2 times a week | 399,540 (82%) | 7,357 (79%) |
| Unknown | 2,676 | 1,379 |
| Non-oily fish |  |  |
| More than 2 times a week | 80,083 (16%) | 1,679 (18%) |
| Less than 2 times a week | 407,698 (84%) | 7,663 (82%) |
| Unknown | 2,333 | 1,367 |
| Fresh fruit |  |  |
| More than 4 servings/day | 37,634 (7.7%) | 902 (9.6%) |
| 2 to 4 servings/day | 277,335 (57%) | 5,036 (54%) |
| < 2 servings/day | 173,269 (35%) | 3,431 (37%) |
| Unknown | 1,876 | 1,340 |
| Cancer | 37,762 (7.7%) | 730 (7.6%) |
| Unknown | 1,610 | 1,140 |
| Diabetes | 25,483 (5.2%) | 840 (8.8%) |
| Unknown | 1,444 | 1,151 |
| Current depression | 110,543 (24%) | 2,325 (28%) |
| Unknown | 21,803 | 2,267 |
| BMI |  |  |
| Normal | 158,816 (33%) | 3,009 (30%) |
| (18.5 kg/m2 to < 25 kg/m2) |  |  |
| Obesity Class I, II or III | 118,867 (25%) | 2,983 (30%) |
| (> 30 kg/m2) |  |  |
| Overweight | 207,399 (43%) | 4,066 (40%) |
| (25 kg/m2 to < 30 kg/m2) |  |  |
| Unknown | 5,032 | 651 |
| Waist circumference | 90.0 (80.0, 99.0) | 92.0 (83.0, 102.0) |
| Unknown | 1,629 | 489 |
| Chronic illness | 155,695 (33%) | 3,731 (41%) |

|  | **Complete baseline social**<br>**isolation<sup>1</sup>**<br>N = 490,114 | **Excluded (missing baseline**<br>**social isolation)<sup>1</sup>**<br>N = 10,709 |
| --- | --- | --- |
| Unknown | 11,688 | 1,610 |
| Non-commute transportation<br>mode |  |  |
| Car-only | 190,673 (39%) | 1,715 (30%) |
| Public transport only | 30,145 (6.2%) | 1,113 (19%) |
| Active transport only | 41,418 (8.5%) | 656 (11%) |
| Mixed-without-car | 33,288 (6.8%) | 731 (13%) |
| Mixed-with-car | 191,461 (39%) | 1,511 (26%) |
| Unknown | 3,129 | 4,983 |
| Commuting transportation mode |  |  |
| Car-only | 164,887 (35%) | 1,112 (16%) |
| Mixed-with-car | 41,615 (8.9%) | 259 (3.8%) |
| Mode excludes car | 54,708 (12%) | 689 (10%) |
| Not employed | 207,033 (44%) | 4,827 (70%) |
| Unknown | 21,871 | 3,822 |
| Driving time |  |  |
| Doesn't drive | 102,922 (21%) | 2,824 (33%) |
| Drives | 379,412 (79%) | 5,771 (67%) |
| Unknown | 7,780 | 2,114 |

<sup>1</sup>n (%); Median (Q1, Q3)
Abbreviations: N = Number of participants; BMI = Body Mass Index; MVPA = Moderate-Vigorous Physical activity, minutes per week; GCSE =
General Certificate of Secondary Education; CSE = Certificate of Secondary Education; NVQ: National Vocational Qualifications; HND: Higher National
Diploma; HNC: Higher National Certificate

**Appendix Table 28.** Characteristics of participants included in main analyses with complete baseline covariates compared to participants excluded due to missing baseline covariate.

|  | Included in main analyses<br>(complete baseline covariates)<sup>1</sup><br>N = 455,072 | Excluded (missing baseline<br>covariates)<sup>1</sup><br>N = 35,042 |
| --- | --- | --- |
| Death | 39,766 (8.7%) | 3,147 (9.0%) |
| Loneliness (Yes) | 82,086 (18%) | 6,733 (20%) |
| Unknown | 7,210 | 944 |
| Age | 58.4 (50.6, 63.8) | 56.9 (49.6, 62.5) |
| Sex (Female) | 250,085 (55%) | 16,882 (48%) |
| Education |  |  |
| O levels/GCSEs or equivalent,<br>CSEs or equivalent | 77,826 (17%) | 4,369 (14%) |
| A levels/AS levels,<br>NVQ/HND/HNC or<br>equivalent | 151,065 (33%) | 9,683 (32%) |
| College or University degree | 147,041 (32%) | 12,309 (40%) |
| None of the above | 79,140 (17%) | 4,080 (13%) |
| Unknown | 0 | 4,601 |
| Household income |  |  |
| Less than £18,000 | 90,325 (20%) | 5,710 (16%) |
| £18,000 to £30,999 | 101,027 (22%) | 6,288 (18%) |
| £31,000 to £51,999 | 102,928 (23%) | 7,086 (20%) |
| Greater than £52,000 | 100,187 (22%) | 8,257 (24%) |
| Do not know/<br>Prefer not to answer | 60,605 (13%) | 7,701 (22%) |
| Unknown | 0 | 0 |
| Live alone | 84,540 (19%) | 6,353 (18%) |
| Unknown | 0 | 0 |
| Ethnicity |  |  |
| Non-white | 22,833 (5.0%) | 2,726 (8.2%) |
| White | 432,239 (95%) | 30,713 (92%) |
| Unknown | 0 | 1,603 |
| Home Population Density |  |  |
| Urban | 387,383 (85%) | 24,982 (83%) |
| Non-urban | 67,689 (15%) | 5,266 (17%) |
| Unknown | 0 | 4,794 |
| Townsend Deprivation Index | -2.2 (-3.7, 0.4) | -1.8 (-3.5, 1.2) |
| Unknown | 0 | 612 |
| Alcohol consumption |  |  |
| Frequent drinking | 197,700 (43%) | 15,612 (45%) |
| Occasional drinking | 169,600 (37%) | 11,699 (33%) |
| Special occasions only | 52,060 (11%) | 4,098 (12%) |
| Never | 35,433 (7.8%) | 3,524 (10%) |
| Unknown | 279 | 109 |
| Smoking |  |  |
| Current | 47,251 (10%) | 4,059 (12%) |
| Never | 248,961 (55%) | 18,614 (54%) |
| Previous | 157,380 (35%) | 12,095 (35%) |
| Unknown | 1,480 | 274 |
| Sleep duration (hours) | 7.0 (6.0, 8.0) | 7.0 (6.0, 8.0) |
| Unknown | 0 | 4 |
| MVPA self-reported | 315.0 (70.0, 630.0) | 245.0 (0.0, 560.0) |
| Unknown | 17,947 | 1,288 |
| Red meat |  |  |
| More than 4 times a week | 225,406 (50%) | 17,457 (50%) |
| 2-4 times a week | 144,493 (32%) | 10,384 (30%) |
| Less than 2 times a week | 84,576 (19%) | 7,100 (20%) |
| Unknown | 597 | 101 |
| Processed meat |  |  |
| More than 2 times a week | 141,015 (31%) | 10,881 (31%) |
| Less than 2 times a week | 313,302 (69%) | 23,961 (69%) |
| Unknown | 755 | 200 |
| Cooked vegetables |  |  |
| More than 4 servings/day | 45,388 (10%) | 4,140 (12%) |
| 2 to 4 servings/day | 322,445 (72%) | 23,895 (70%) |
| < 2 servings/day | 82,208 (18%) | 6,236 (18%) |
| Unknown | 5,031 | 771 |
| Oily fish |  |  |
| More than 2 times a week | 81,251 (18%) | 6,647 (19%) |
| Less than 2 times a week | 371,524 (82%) | 28,016 (81%) |
| Unknown | 2,297 | 379 |
| Non-oily fish |  |  |
| More than 2 times a week | 74,239 (16%) | 5,844 (17%) |
| Less than 2 times a week | 378,870 (84%) | 28,828 (83%) |
| Unknown | 1,963 | 370 |
| Fresh fruit |  |  |
| More than 4 servings/day | 35,006 (7.7%) | 2,628 (7.6%) |
| 2 to 4 servings/day | 258,880 (57%) | 18,455 (53%) |
| < 2 servings/day | 159,632 (35%) | 13,637 (39%) |
| Unknown | 1,554 | 322 |
| Cancer | 35,272 (7.8%) | 2,490 (7.2%) |
| Unknown | 1,368 | 242 |
| Diabetes | 23,368 (5.1%) | 2,115 (6.1%) |
| Unknown | 1,193 | 251 |
| Current depression | 102,243 (23%) | 8,300 (25%) |
| Unknown | 19,680 | 2,123 |
| BMI |  |  |
| Normal<br>(18.5 kg/m<sup>2</sup> to < 25 kg/m<sup>2</sup>) | 147,944 (33%) | 10,872 (32%) |
| Obesity Class I, II or III<br>(≥ 30 kg/m<sup>2</sup>) | 109,956 (24%) | 8,911 (26%) |
| Overweight<br>(25 kg/m<sup>2</sup> to < 30 kg/m<sup>2</sup>) | 192,908 (43%) | 14,491 (42%) |
| Unknown | 4,264 | 768 |
| Waist circumference | 90.0 (80.0, 99.0) | 91.0 (82.0, 100.0) |
| Unknown | 1,273 | 356 |
| Chronic illness | 144,076 (32%) | 11,619 (34%) |
| Unknown | 10,610 | 1,078 |
| Non-commute transportation<br>mode |  |  |
| Car-only | 178,134 (39%) | 12,539 (39%) |
| Public transport only | 28,280 (6.2%) | 1,865 (5.8%) |
| Active transport only | 38,245 (8.4%) | 3,173 (9.9%) |
| Mixed-without-car | 31,225 (6.9%) | 2,063 (6.5%) |
| Mixed-with-car | 179,188 (39%) | 12,273 (38%) |
| Unknown | 0 | 3,129 |
| Commuting transportation mode |  |  |
| Car-only | 160,961 (35%) | 3,926 (30%) |
| Mixed-with-car | 40,807 (9.0%) | 808 (6.1%) |
| Mode excludes car | 53,208 (12%) | 1,500 (11%) |
| Not employed | 200,096 (44%) | 6,937 (53%) |
| Unknown | 0 | 21,871 |
| Driving time |  |  |
| Doesn't drive | 94,871 (21%) | 8,051 (24%) |
| Drives | 353,489 (79%) | 25,923 (76%) |
| Unknown | 6,712 | 1,068 |

<sup>1</sup>n (%); Median (Q1, Q3)
Abbreviations: N = Number of participants; BMI = Body Mass Index; MVPA = Moderate-Vigorous Physical activity, minutes per week; GCSE =
General Certificate of Secondary Education; CSE = Certificate of Secondary Education; NVQ: National Vocational Qualifications; HND: Higher National
Diploma; HNC: Higher National Certificate

**Appendix Text 3.** STROBE Checklist STROBE Statement—Checklist of items that should be included in reports of cohort studies.

|  | Item No | Recommendation | Page No |
| --- | --- | --- | --- |
| **Title and abstract** | 1 | (a) Indicate the study's design with a commonly used term in the title or the abstract<br>(b) Provide in the abstract an informative and balanced summary of what was done and what was found | p. 1<br>pp. 1-2 |
| **Introduction** |  |  |  |
| Background/rationale | 2 | Explain the scientific background and rationale for the investigation being reported | pp. 2-3 |
| Objectives | 3 | State specific objectives, including any prespecified hypotheses | p. 3 |
| **Methods** |  |  |  |
| Study design | 4 | Present key elements of study design early in the paper | pp. 4-5 |
| Setting | 5 | Describe the setting, locations, and relevant dates, including periods of recruitment, exposure, follow-up, and data collection | p. 4 |
| Participants | 6 | (a) Give the eligibility covariates, and the sources and methods of selection of participants. Describe methods of follow-up<br>(b) For matched studies, give matching covariates and number of exposed and unexposed | pp. 4-5<br>NA |
| Variables | 7 | Clearly define all outcomes, exposures, predictors, potential confounders, and effect modifiers. Give diagnostic covariates, if applicable | pp. 4-5 |
| Data sources/<br>measurement | 8* | For each variable of interest, give sources of data and details of methods of assessment (measurement). Describe comparability of assessment methods if there is more than one group | pp. 4-5 |
| Bias | 9 | Describe any efforts to address potential sources of bias | pp. 5-7, 13-14 |
| Study size | 10 | Explain how the study size was arrived at | p. 4, Appendix Figure 1 |
| Quantitative variables | 11 | Explain how quantitative variables were handled in the analyses. If applicable, describe which groupings were chosen and why | p. 4-5, Appendix Text 1 |
| Statistical methods | 12 | (a) Describe all statistical methods, including those used to control for confounding<br>(b) Describe any methods used to examine subgroups and interactions<br>(c) Explain how missing data were addressed<br>(d) If applicable, explain how loss to follow-up was addressed<br>(e) Describe any sensitivity analyses | pp. 5-6<br>pp. 5-6<br>p. 6-7<br>p. 13<br>pp. 6-7 |
| **Results** |  |  |  |
| Participants | 13* | (a) Report numbers of individuals at each stage of study—eg numbers potentially eligible, examined for eligibility, confirmed eligible, included in the study, completing follow-up, and analysed | p. 4, Appendix Figure 1 |
|  |  | (b) Give reasons for non-participation at each stage | Appendix Figure 1 |
|  |  | (c) Consider use of a flow diagram | Appendix Figure 1 |
| Descriptive data | 14* | (a) Give characteristics of study participants (eg demographic, clinical, social) and information on exposures and potential confounders<br>(b) Indicate number of participants with missing data for each variable of interest<br>(c) Summarise follow-up time (eg, average and total amount) | p.7, Table 1<br>Table 1, Appendix Tables 27-28<br>pp. 4, 7 |
| Mortality outcome data | 15* | Report numbers of outcome events or summary measures over time | p.7, Table 1 |
| Main results | 16 | (a) Give unadjusted estimates and, if applicable, confounder-adjusted estimates and their precision (eg, 95% confidence interval). Make clear which confounders were adjusted for and why they were included<br>(b) Report category boundaries when continuous variables were categorized<br>(c) If relevant, consider translating estimates of relative risk into absolute risk for a meaningful time period | pp. 8-11<br>Appendix Figure 2-3 and Text 1<br>Appendix Text 1<br>p. 9 |
| Other analyses | 17 | Report other analyses done—eg analyses of subgroups and interactions, and sensitivity analyses | pp. 9-11 |
| **Discussion** |  |  |  |
| Key results | 18 | Summarise key results with reference to study objectives | pp. 11-12 |
| Limitations | 19 | Discuss limitations of the study, taking into account sources of potential bias or imprecision. Discuss both direction and magnitude of any potential bias | pp. 12-14 |
| Interpretation | 20 | Give a cautious overall interpretation of results considering objectives, limitations, multiplicity of analyses, results from similar studies, and other relevant evidence | pp. 11-12, 14 |
| Generalisability | 21 | Discuss the generalisability (external validity) of the study results | p. 12-13 |
| **Other information** |  |  |  |
| Funding | 22 | Give the source of funding and the role of the funders for the present study and, if applicable, for the original study on which the present article is based | p. 14 |

\*Give information separately for exposed and unexposed groups.
**Note:** An Explanation and Elaboration article discusses each checklist item and gives methodological background and published examples of transparent reporting. The STROBE checklist is best used in conjunction with this article (freely available on the Web sites of PLoS Medicine at http://www.plosmedicine.org/, Annals of Internal Medicine at http://www.annals.org/, and Epidemiology at http://www.epidem.com/). Information on the STROBE Initiative is available at http://www.strobe-statement.org.

